# Grandfathers and Grandsons: Social Security Expansion and Child Health in China

**DOI:** 10.1101/2022.04.23.22274222

**Authors:** Jinyang Yang, Xi Chen

**Affiliations:** School of Economics, Huazhong University of Science and Technology; Department of Agricultural and Applied Economics, Virginia Tech. 250 Drillfield Dr, Blacksburg, VA 24060; Department of Health Policy and Management, Department of Economics, Yale University; IZA - Institute of Labor Economics. 60 College St, New Haven, CT 06520

**Keywords:** Social pension, Child health, Inter-generational relationship, Intra-household allocation, Migration, Living arrangement, China, H23, H31, H55, I38, J22, O15

## Abstract

We examine the multigenerational impacts of a nationwide social pension program in China, the New Rural Pension Scheme (NRPS). NRPS was rolled out in full scale since 2012, and rural enrollees over age 60 are eligible to receive a minimum of 70 CNY non-contributory monthly pension. We leverage age eligibility and variations in pension receipt to identify the inter-generational effect of NRPS on health among grandchildren. We find NRPS substantially increases child weight without impacting height. Overall, child BMI z score increases by 1.09, which is largely driven by grandfathers’ pension receipt raising rates of overweight and obesity among grandsons. Among the potential mechanisms, our findings are more plausibly explained by a mixture of income effect, son preference, and rising inter-generational co-residence and childcare.

## 1 Introduction

Children living in rural areas in developing countries are more likely undernourished than their counterparts in urban areas or developed countries. In recent years, the double burden of child malnutrition, characterized by the coexistence of nutritional insufficiency and nutritional imbalance (e.g. overweight or obesity), has also become prevalent (Wells et al., 2020). Similarly, a salient gap in child nutritional status persists between rural and urban China, and existing studies document the prevalence and growth of rural child obesity (Piernas et al., 2015; Song et al., 2015) and anemia (Zhang et al., 2013). As an increasing share of children in rural China are taken care by grandparents while their parents devote more time to labor market, it is important to understand the growing importance of grandparents in shaping child nutritional status and, more generally, human capital development.

Cash transfers offer a viable way to redistribute resources and address child nutritional disadvantages, which help improve health, education, and labor market outcomes in adulthood (Duflo, 2003; Aizer et al., 2016). Cash transfers are likely more efficient when the targeted population is also the main decision maker, therefore recipients fully internalize the returns to investment. While a large body of literature has shed light on cash transfers to parents and child health, less is known about multi-generational impacts of cash transfers. It is also under-explored whether the influence of grandparents on child health shows any gendered pattern. The answer to these questions may have policy implications for China and other developing countries where multi-generational co-residence or decision-making is common.

This paper evaluates the impacts of the world’s largest social pension program that benefits hundreds of millions of rural residents, the New Rural Pension Scheme (NRPS), on grandchildren’s nutrition status in China. Previous studies have leveraged another social pension program - the Old Age Pension (OAP) in South Africa - to understand the inter-generational health effects. Both OAP and NRPS took a few years to roll out to all areas, but they differ in two key aspects: the size of OAP payment to beneficiaries is more than twice the median per capita income of rural South Africans, while NRPS payment accounts for about 10 percent of per capita income in China; the eligibility for OAP is means tested, in contrast, all residents with rural hukou ^1^ are eligible to enroll in NRPS.

The NRPS started county-by-county roll-out in 2009, and by the end of 2012 all counties had been covered. The universal NRPS eligibility criteria (rural residents age over 60) allow us to employ a quasi-experimental design to identify the multigenerational effects of NRPS. We focus on rural children age under 12 years (6-144 months) and compile a sample of their households. We use household age eligibility as an instrument for NRPS pension receipt, in addition to controlling for child and household characteristics, cohort effects, county and interview year fixed effects.

Our findings reveal that pension receipt substantially changes grandchildren’s short-term nutrition status, as measured by their increased BMI z score, overweight or obesity, but no reduced underweight. The effect has not been manifested in longer term outcomes, such as height. Moreover, we show gendered pattern for pension recipients. Specifically, grandfathers’ pension receipt has both economically and statistically significant effect on grandsons’ weight, while the impact on granddaughters are statistically insignificant. In contrast, we observe no effect of grandmothers receiving pension on grandchildren.

We examine a number of potential mechanisms. Household income increases by more than 10% on average around NRPS eligible age. When examining the impacts by income sources, we show the change in pubic transfers (NRPS included) is the main contributor. While co-residence arrangement does not change with NRPS pension receipt, our data suggest that over time grandparents become more likely to be a main caregiver for children under age 12, and mothers have a declined rate of serving this role.^2^ Moreover, adult child migration increases slightly. It is plausible that NRPS changes child weight through the channel of income expansion and nutrients intake, and grandparents allocate more time to child care though they are less knowledgeable about scientifically feeding children. When differentiating father’s parents and mother’s parents, father’s father imposes the most salient effect on grandsons’ health, suggesting that son preference may enhance the impact on boys.

This study attempts to make three main contributions to the literature. Firstly, it sheds light on the role of grandparents in grandchildren’s human capital formation. More specifically, it is, to our knowledge, the first paper that explores multigenerational effects of NRPS on grandchildren’s nutritional outcomes. Existing studies have evaluated a comprehensive set of NRPS impacts, including on elderly labor supply (Ning et al., 2016; Huang and Zhang, 2021), intra-household transfers (Huang and Zhang, 2021; Chen, Eggleston and Sun, 2017; Nikolov and Adelman, 2019), senior health (Cheng et al., 2018a; Chen, Wang and Busch, 2019; Huang and Zhang, 2021), healthcare utilization (Chen, Eggleston and Sun, 2017), living arrangement with adult children (Chen, Eggleston and Sun, 2017; Eggleston, Sun and Zhan, 2018; Cheng et al., 2018b) and adult child migration (Eggleston, Sun and Zhan, 2018). However, few examine the multi-generational effects on grandchildren except Huang and Zhang (2021) that investigate grandchildren’s self-reported health. Similar evaluations have been conducted for South Africa’s OAP (Case and Deaton, 1998; Case, 2001; Duflo, 2003; Maitra and Ray, 2003; Jensen, 2004), in which Duflo (2003) shows that only grandmothers’ OAP receipt has significant impacts on granddaughters’ weight and height. Given the differences between NRPS and OAP as well as much higher fertility rate in South Africa than in China, their family decision-making on time and resource allocated to children may vary.

Secondly, this paper adds to the studies on intra-household resource allocation. Empirical studies have focused on exogenous income or wealth shocks to household members. A growing literature suggests that economic resources in the hands of women are spent more on nutrition to improve child health than are resources in the hands of men (Duflo, 2012; Duflo and Udry, 2004; Duflo, 2003; Rangel, 2006; Lundberg, 2005; Dizon-Ross and Jayachandran, 2022). This paper, instead, shows the salient impact of men’s permanent income change on worsening child health. We lend further support to this idea that females’ relative empowerment in the family context may promote child health in future generations. Since families in which women own more economic resources could differ in many respects from families in which women have no access to such resources, our context of unconditional universal pension income above an age cut-off should mitigate this bias.

Thirdly, this study may relate to the literature on the unintended consequences of policies or family arrangements on child obesity. Studies in developed countries, such as the United States, find food assistance programs, originally designed to relieve hunger and under-nutrition, unintentionally increase child obesity (See the review by Cawley (2015)). Fewer evidence are from developing countries. In China, co-residence with grandparents may increase grandchildren’s weight, and the effects are stronger in rural areas (He, Li and Wang, 2018).

This study distinguishes from and may advance Duflo (2003), i.e., the study closest to ours, in two main aspects. First, the positive income shock we leverage is universal to all rural elderly in China above age 60, not just limited to older adults in some disadvantaged groups. This universal eligibility of NRPS may eliminate the concern over endogenous take-up decisions. While Duflo (2003) overcomes endogeneity of pension enrollment by using age eligibility as instruments, the means-tested feature of OAP in South Africa determines that unobservables correlated with pre-treated household income and health outcomes might also be correlated with demographic structure and thus the presence of eligible household member, which may invalidate the instruments. Second, instead of using cross-sectional data, we use a nationally representative sample that follows up household members and their descendants in three waves, one before the full roll-out of NRPS and two afterwards. We further compare child outcomes of eligible and ineligible households, before and after the expansion of NRPS.

The rest of the paper is organized as follows. Section 2 introduces the backgrounds of rural child care, migration and the expansion of NRPS. In Section 3 we describe our data, and in Section 4 we present the empirical strategy. Section 5 presents the estimation results of NRPS on child weight and height. In Section 6 we explore potential mechanisms and other related outcomes. We conclude in Section 7.

## 2 Child Care, Migration, and Social Security Expansion in Rural China

The New Rural Pension Scheme (NRPS) is a nationwide social pension program that aims to enroll rural population in China. The NRPS pilot was launched in 320 out of 2,853 counties in 2009, reached 838 counties by 2010. The pace of NRPS rollout in 2009-2010 was moderate, followed by more rapid expansion since 2011, which ended up covering all counties by the end of 2012. The participation rate at the individual level rose dramatically: for NRPS eligible older adults, only 3 percent received pension by 2010, but the number increased to above 40 percent by 2012, according to our tabulation of CFPS data.^3^ NRPS now consists the most important safety net for older adults in rural China.

NRPS was designed to incorporate two parts, a non-contributory social pension benefit and a voluntary defined-contribution pension savings scheme. Residents with a rural registration (hukou type) are all eligible to enroll in this program. Specifically, enrollees over age 60 are eligible to receive a non-contributory pension set by the central government (a minimum 55 Chinese yuan, about 8 US dollars) per month per person in 2009, which increased to at least 70 Chinese yuan in 2014. While most counties adopt the lower-bound pension benefit, the size of the benefit can be raised by local government, depending on their fiscal revenues. Enrollees under 60 must contribute a minimum annual premium of 100 Chinese yuan to their individual account, which is matched with at least additional 30 yuan from local government.

In 2020, 285.6 million rural residents work in urban sectors.^4^ A large proportion of this population live separately from their children.^5^ Consequently, an increasingly sizable proportion of rural children live with or cared by their grandparents in China. In Chinese culture, multi-generational co-residence is esteemed as a symbol of filial piety and family harmony (*Tian Lun Zhi Le*). In 2012-2014, around 45% of elderly at age 60 in the China Family Panel Studies (CFPS), a national representative longitudinal survey, co-reside with children under 12, reaching its highest level across all ages of older adults (Figure A1).

The share of grandparents taking the role as the primary caregiver for grandchildren also rises over time, at least partly due to more job opportunities for parents to migrate to work and lack of childcare provision in rural areas. In CFPS 2012-2014, over 30 percent of rural children under 12 had grandparents as their primary daytime caregivers, and slightly below 30 percent had grandparents as primary nighttime caregivers (see Figure 1).

**Figure 1:**
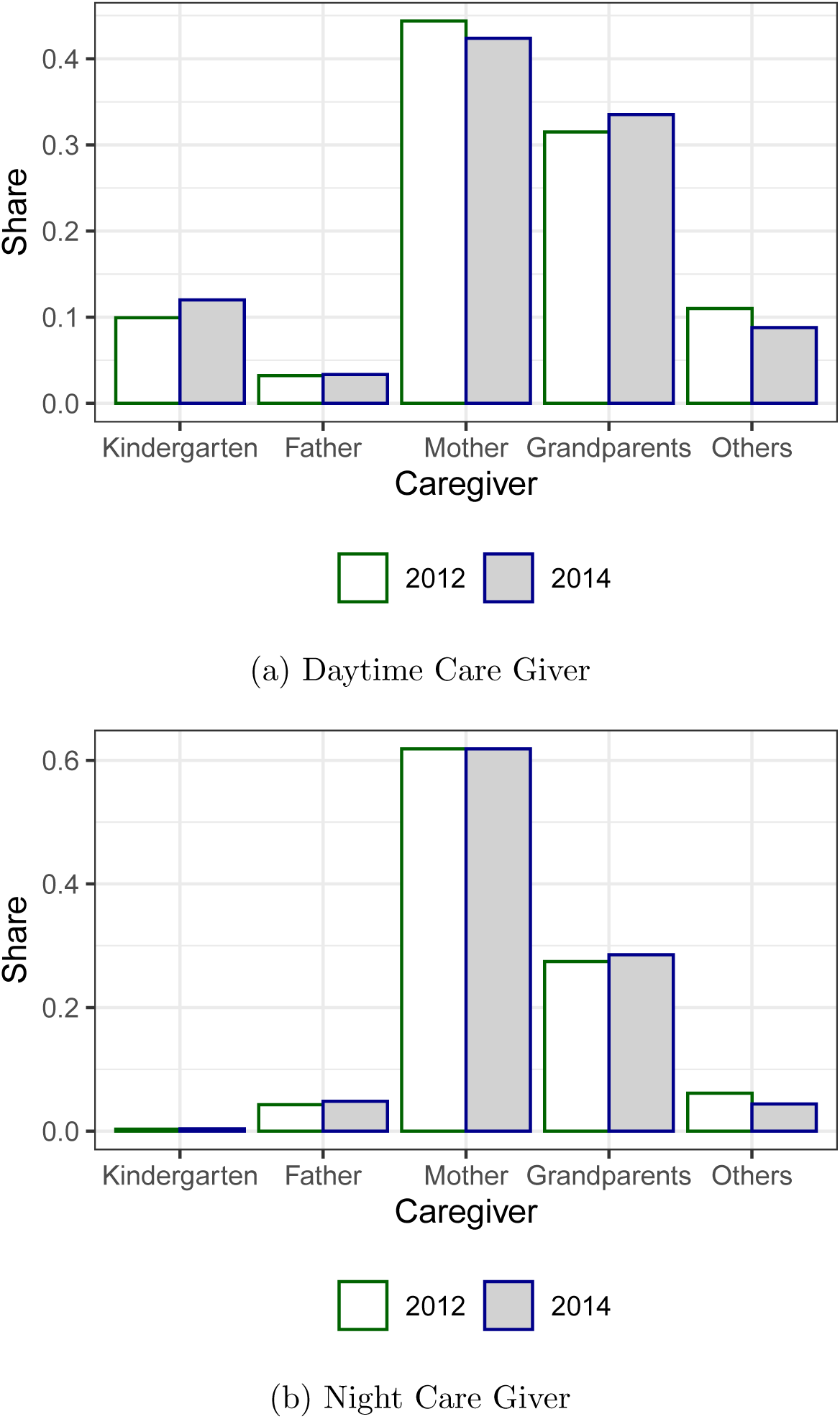
Share of Primary Caregivers for Children under Age 12. *Note:* Authors’ tabulations of China Family Panel Studies wave 2012 and 2014 data. Daytime childcare giver is defined by CFPS question ”who is usually the main care giver of the child during the daytime?” Night childcare giver is defined by CFPS question ”who is usually the main care giver of the child during the night?” Wave 2010 is not included because the survey question in wave 2010 is different from that in wave 2012 and 2014. It only asks who mainly take care of the children, regardless of daytime and night.

Given the fact that NRPS accounts for a main income source for grandparents, this public transfer aiming at the rural elderly are likely spent on grandchildren, such as through more food intake, with some unintended nutritional consequences (He, Li and Wang, 2018). Firstly, a majority of grandparents in countryside are illiterate or semi-illiterate. Their famine experiences in early life and limited knowledge on child care may determine that securing adequate food is at the core of their child rearing. They might possess biased view of healthy diet and physical activities. For instance, high starch food like rice and high fat meat like pork belly are favored in many rural households. “Chubby Boy” (*Da Pang Xiao Zi*) is considered healthy. Secondly, informal labor participation rate of the elderly is higher in countryside. Unlike their urban counterparts, rural residents do not have statutory retirement age, neither do they have pension support before NRPS was introduced. The lack of young labor in agricultural sector due to migration often requires grandparents to farm in older ages while taking care of grandchildren. Grandchildren could be left unattended in busy season. Thirdly, as part of Chinese culture, grandparents tend to spoil their grandchildren, especially grandsons. Requests made by children, such as extra pocket money for snacks, are more likely satisfied by grandparents than by parents. A recent study by Silverstein and Zhang (2020) finds financial transfers from grandparents to grandchildren often follow a male lineage, and is the greatest to grandson-only families in which parents are first-born sons.

## 3 Data and Descriptive Statistics

### 3.1 Data

This study compares anthropometric status of children in households receiving NRPS to those in households without. The data were compiled from the China Family Panel Studies (CFPS), a nationwide biennial survey of Chinese households conducted by Peking University since 2010. It covers 25 provinces and is representative of 95 percent of China’s total population. Its 2010 baseline survey constitutes of 14,960 households and 42,590 individuals. Core household members and members of their newly formed families were permanently followed up in the subsequent waves. The 2012 and 2014 waves respectively surveyed 6,453 and 6,608 children under 12. 35,719 and 37,147 adults were surveyed in 2012 and 2014, respectively, with 9,130 and 9,934 individuals age over 60.

Height and weight of children under 12 are all reported by their main caregivers. For each age in months, we use BMI z scores to measure short-run child nutrition and height-for-age z scores to measure long-run child nutrition. ^6^

CFPS asks each adult whether they have received NRPS and, if yes, what year and month they started to receive. Household pension receipt is coded as whether there is an adult receiving NRPS, and household eligibility is coded as whether an adult ages over 60.

Our study focuses on children under 12 as they often demand more intensive care, and nutrition in younger ages could have persistent impacts on adulthood. They are also suffering from double burden of malnutrition. As shown in Figure 2, while rural children at all ages under 12 are shorter than their urban counterparts, they surpass urban children in body weight. To study the role of NRPS, we compiled CFPS waves 2012 and 2014, and matched children under 12 years (6-144 months) with characteristics of households and their members. Our sample criteria include: 1) children with rural hukou; 2) excluding children age 0-5 months, due to concerns over measurement error; and 3) excluding children with BMI z scores or height-for-age z scores in the top or bottom 1 percentiles. Finally, we obtain a sample of 3,898 boys and 3,468 girls. Wave 2010 is excluded because by then only up to 56 out of 162 counties in CFPS were covered by NRPS.

**Figure 2:**
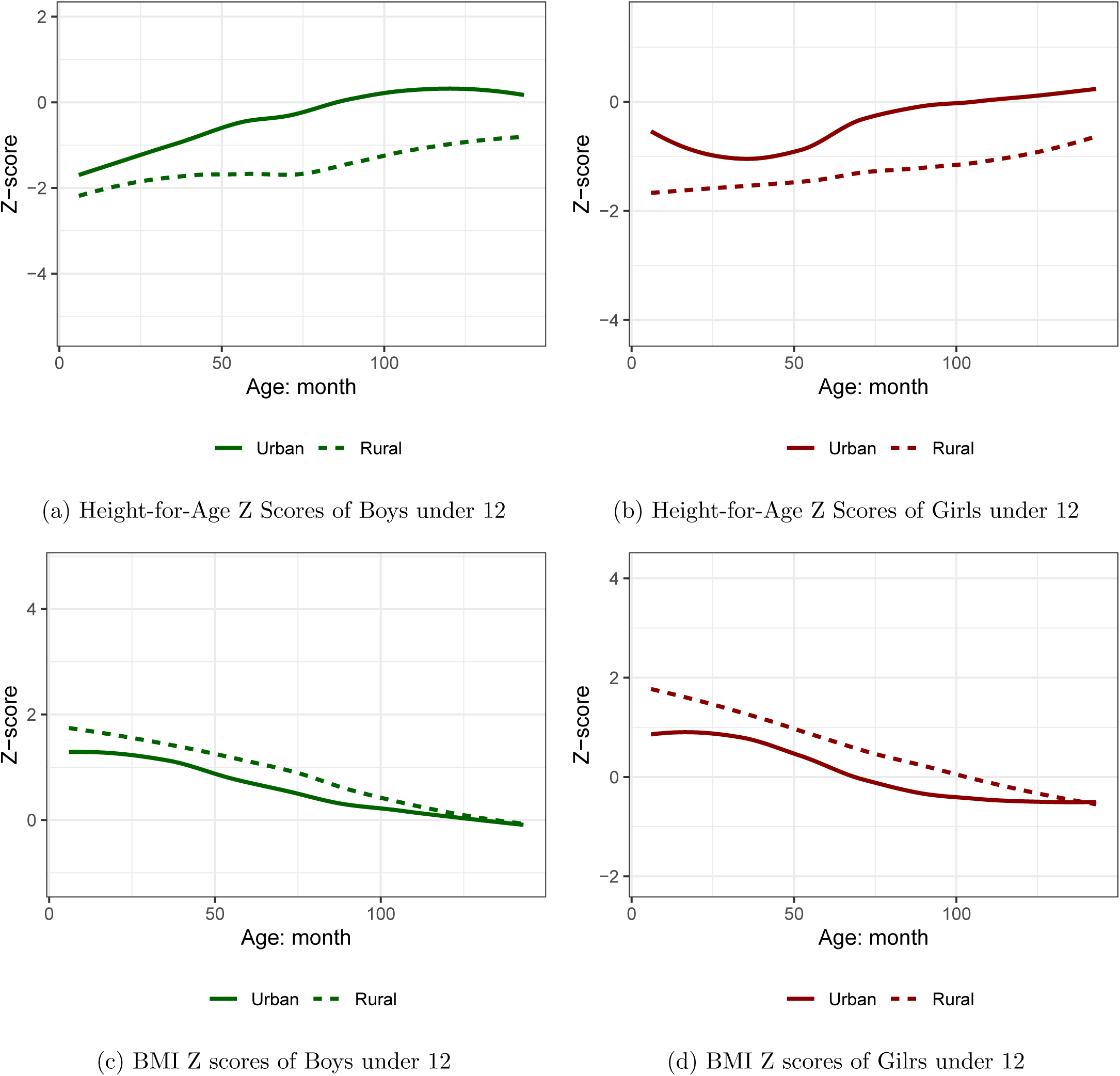
Nutritional Status of Rural and Urban Children under Age 12. *Note:*Authors’ tabulations of China Family Panel Studies wave 2010, 2012 and 2014 data. This figure present local regression of height-for-age z score and BMI z score on child age (months). Height-for-age z scores and BMI z scores are calculated based on Child Growth Standards (0-5 years) and Growth Reference (5-19 years) developed by World Health Organization (WHO). Children age less than 6 months are dropped.

### 3.2 Descriptive Statistics

Table 1 reports summary statistics for samples classified by NRPS pension receipt among household members. Panel A shows age, gender and anthropometric outcomes of children. Underweight is defined as BMI z score less than -2; overweight is defined as BMI z score greater than 2; obesity is defined as BMI z score greater than 3. We also define stunting by height-for-age z scores below -2.

**Table 1:** Summary Statistics by Gender of Pension Receipt.

|  | Either | Male | Female | None | T-test<br>p-values |
| --- | --- | --- | --- | --- | --- |
| **Panel A: Children** |  |  |  |  |  |
| Gender (boy=1) | 0.513<br>(0.500) | 0.518<br>(0.500) | 0.511<br>(0.500) | 0.540<br>(0.498) | 0.075 |
| Age (month) | 75.040<br>(40.352) | 75.643<br>(39.924) | 76.878<br>(40.890) | 68.921<br>(40.150) | 0.000 |
| BMI z score | 1.454<br>(3.730) | 1.269<br>(3.491) | 1.491<br>(3.760) | 1.303<br>(3.575) | 0.164 |
| Obese (yes=1) | 0.196<br>(0.397) | 0.170<br>(0.376) | 0.204<br>(0.403) | 0.195<br>(0.396) | 0.923 |
| Overweight (yes=1) | 0.330<br>(0.470) | 0.308<br>(0.462) | 0.331<br>(0.471) | 0.309<br>(0.462) | 0.130 |
| Underweight (yes=1) | 0.105<br>(0.307) | 0.103<br>(0.303) | 0.101<br>(0.301) | 0.116<br>(0.320) | 0.248 |
| Height-for-age z score | -1.843<br>(3.080) | -1.598<br>(2.878) | -1.956<br>(3.134) | -1.679<br>(2.909) | 0.067 |
| Stunting (yes=1) | 0.403<br>(0.490) | 0.383<br>(0.486) | 0.404<br>(0.491) | 0.367<br>(0.482) | 0.012 |
| **Panel B: Household** |  |  |  |  |  |
| Inc per capita (1000 yuan) | 6.770<br>(5.805) | 6.770<br>(5.752) | 6.539<br>(5.486) | 7.447<br>(6.907) | 0.000 |
| Household size | 6.319<br>(1.857) | 6.294<br>(1.662) | 6.489<br>(1.992) | 5.287<br>(1.789) | 0.000 |
| Father's age (year) | 35.051<br>(5.414) | 34.817<br>(5.153) | 35.490<br>(5.517) | 33.827<br>(6.460) | 0.000 |
| Mother's age (year) | 32.804<br>(5.591) | 32.623<br>(5.381) | 33.168<br>(5.683) | 32.002<br>(6.296) | 0.000 |
| Father's edu (year) | 8.129<br>(3.183) | 8.196<br>(3.062) | 7.990<br>(3.291) | 8.131<br>(3.436) | 0.983 |
| Mother's edu (year) | 7.059<br>(3.688) | 7.243<br>(3.627) | 6.932<br>(3.704) | 7.316<br>(3.834) | 0.018 |
| Urban (yes=1) | 0.294<br>(0.455) | 0.252<br>(0.434) | 0.296<br>(0.457) | 0.353<br>(0.478) | 0.000 |
| *# of members in age group:* |  |  |  |  |  |
| < 5 | 0.855<br>(0.864) | 0.800<br>(0.768) | 0.832<br>(0.906) | 0.913<br>(0.830) | 0.021 |
| 6 – 15 | 1.201<br>(1.013) | 1.203<br>(1.025) | 1.267<br>(1.012) | 0.950<br>(0.873) | 0.000 |
| 16 – 24 | 0.301<br>(0.600) | 0.267<br>(0.555) | 0.348<br>(0.644) | 0.365<br>(0.678) | 0.000 |
| 25 – 49 | 2.152<br>(0.951) | 2.078<br>(0.911) | 2.224<br>(1.015) | 2.126<br>(0.857) | 0.340 |
| *Presence of household member:* |  |  |  |  |  |
| Male age over 50 (yes=1) | 0.784<br>(0.411) | 0.997<br>(0.054) | 0.685<br>(0.465) | 0.439<br>(0.496) | 0.000 |
| Female age over 50 (yes=1) | 0.901<br>(0.299) | 0.847<br>(0.360) | 0.988<br>(0.110) | 0.453<br>(0.498) | 0.000 |
| Male age over 70 (yes=1) | 0.182<br>(0.386) | 0.249<br>(0.432) | 0.177<br>(0.381) | 0.043<br>(0.202) | 0.000 |
| Female age over 70 (yes=1) | 0.236<br>(0.424) | 0.128<br>(0.334) | 0.319<br>(0.466) | 0.062<br>(0.241) | 0.000 |
| Male pensioner (yes=1) | 0.604<br>(0.489) | 1.000<br>(0.000) | 0.422<br>(0.494) | 0.000<br>(0.000) |  |
| Female pensioner (yes=1) | 0.684<br>(0.465) | 0.478<br>(0.500) | 1.000<br>(0.000) | 0.000<br>(0.000) |  |
| Observations | 1445 | 889 | 990 | 5921 |  |

*Notes:* This sample comes from CFPS wave 2012 and 2014. “Either” includes households that have male or female pension receipt. “Male” pension receipt status includes households that have male or both gender receipt. “Female” pension receipt status includes households that have female or both gender receipt. The last column reports t-test p-values of households with and without pension receipt. Standard deviations are in parentheses.

On the one hand, children in NRPS recipient households have larger BMI but shorter stature. More specifically, children in households with female pension recipient have the largest average BMI z score among all groups, as well as the highest overweight and obesity rates. However, average child BMI z score in households with male pension recipient is similar to that in households without pension recipient, so are the overweight, obesity and underweight rates. Children in recipient households are also more disadvantageous in height and stunting rate. Though somewhat different, the gaps of child weight and height between households with and without pension recipients are statistically insignificant in most outcomes using a two sample t test (see the last column of Table 1).

On the other hand, children in households with and without NRPS recipients are heterogeneous in age, gender and household backgrounds.All of them show statistically difference as well. Firstly, average age of children in households with NRPS receipt are 6 months older than those without. Secondly, households receiving NRPS tend to have larger household size, lower income per capita, and larger chance of residing in rural areas than their non-receipt counterparts.^7^ To understand whether the differences in child nutrition can be partially attributable to NRPS, we next adopt an instrumental variable approach while controlling for a rich set of child and household characteristics.

## 4 Estimation Strategy

The impact of receiving pension by grandparents on the health outcomes of grand-children can be identified by the models below:

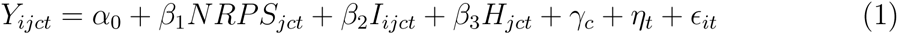

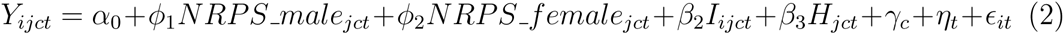

where *Y_ijct_* measures the health outcomes (BMI z scores or height-for-age z scores) of child *i* in household *j* county *c* at time *t*. In equation (1), *NRPS_jct_* is a dummy indicating whether there is an adult receiving NRPS pension in household *j*, county *c* and year *t*. When separating the effects of NRPS by male and female pensioners, we replace *NRPS_jct_* with two dummies, *NRPS_male_jct_* and *NRPS_female_jct_* in equation (2), which indicates whether there is a male or female pensioner in household *j*, county *c* and year *t*. *I_ijct_* includes a set of dummies for individual characteristics, such as age (in months) and gender. *H_jct_* controls for household observable characteristics, such as household size, urban or rural residence, father’s and mother’s year of education and age, number of members ages 0-5, 6-15, 16-24, 25-49, and dummy variables respectively indicating the presence of a woman age over 50, and a man age over 50. For robustness checks, we also control for the presence of males and females over age 70 in the household, respectively. *γ_c_* and *η_t_* are county and time fixed effects, respectively.

The decision to take up NRPS is not random. For instance, the elderly who receive less support from adult children tend to miss the opportunity of enrolling in the program (Chen, Hu and Sindelar, 2020), and their grandchildren may have worse nutrition outcomes if parents live away from home village and offer less support to child care. Fortunately, the exogenously determined age eligibility for NRPS pension receipt offers us an instrument to address this endogeneity. Due to this policy, there is a discontinuity in pension receipt at age 60 (see Figure 3a). In equation (1), we use a dummy variable indicating the existence of an eligible household member as an instrument for *NRPS_jct_*. In equation (2), two variables, male and female pension eligibility statuses are used as instruments for *NRPS_male_jct_* and *NRPS_female_jct_*, separately.

**Figure 3:**
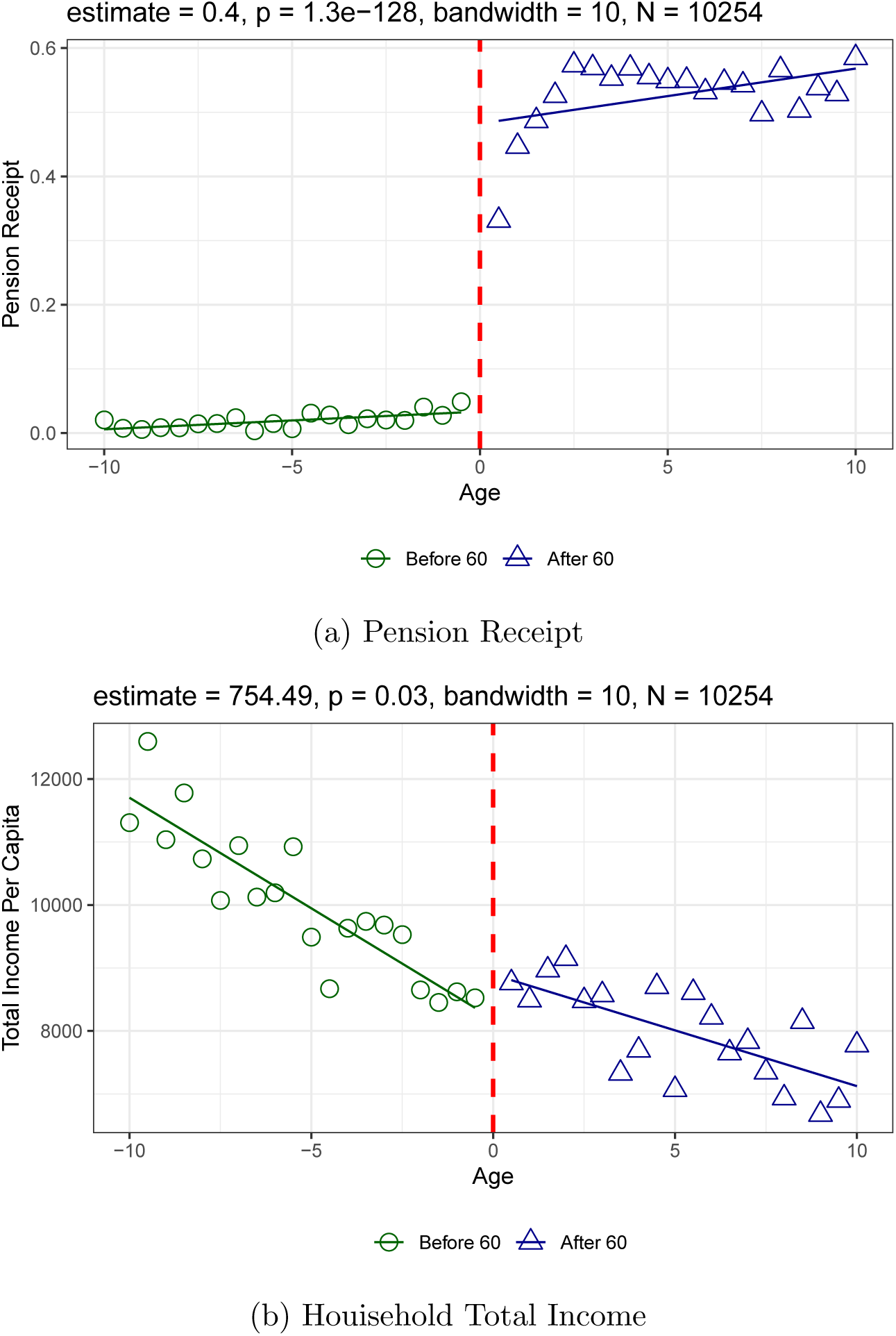
Effects of NRPS on Household Income. *Note*: The data comes from CFPS wave 2012 and 2014. Panel (a) shows the change of pension receipt rate by adult’s age (in 0.5 years). Panels (b) displays the average household income per capita at ages (in 1.5 years) 50-70. Ages are centered at 60. Household total income per capita is calcuated by household gross income divided by number of household members. The solid lines show the fitted lines before and after 60 years old. The RD estimation results are displayed in each panel using adult observations.

## 5 Results

### 5.1 Effects of Age Eligibility on Pension Receipt

In this section we present the first-stage regression results of household pension receipt on age eligibility. The results will confirm if pension age eligibility in the 2SLS estimations provides us strong instruments. In all regression tables, we cluster standard errors at the county level, unless stated otherwise. We also use CFPS sample weights in all estimations.

Table 2 shows the full sample results. Columns 1-4 report the results without distinguishing gender of the pension recipient. Column 3 presents results from our baseline model. We incrementally control for child and household characteristics, and a set of fixed effects. The estimates on NRPS eligibility are very stable across specifications. Overall, household age eligibility significantly increases likelihood of pension receipt by 45-47 percentage points. The F-statistic for the excluded instrument demonstrates that age eligibility is a strong instrument for household pension receipt.

**Table 2:** First Stage Regressions: Full Sample Results.

|  | Either gender receipt |  |  |  | Male | Female | Male | Female |
| --- | --- | --- | --- | --- | --- | --- | --- | --- |
|  | (1) | (2) | (3) | (4) | (5) | (6) | (7) | (8) |
| NRPS eligibility | 0.473***<br>(0.027) | 0.448***<br>(0.029) | 0.454***<br>(0.029) | 0.455***<br>(0.029) |  |  |  |  |
| Male eligibility |  |  |  |  | 0.400***<br>(0.031) | 0.050**<br>(0.020) | 0.402***<br>(0.032) | 0.039*<br>(0.021) |
| Female eligibility |  |  |  |  | 0.038*<br>(0.020) | 0.404***<br>(0.026) | 0.047**<br>(0.023) | 0.412***<br>(0.027) |
| Age and gender | Y | Y | Y | Y | Y | Y | Y | Y |
| Household covariates |  | Y | Y | Y | Y | Y | Y | Y |
| County FEs |  |  | Y | Y | Y | Y | Y | Y |
| Year FEs | Y | Y | Y | Y | Y | Y | Y | Y |
| Older seniors |  |  |  | Y |  |  | Y | Y |
| F statistics | 5376.41 | 238.65 | 245.08 | 246.17 | 166.49 | 241.44 | 157.82 | 232.84 |
| R<sup>2</sup> | 0.361 | 0.365 | 0.436 | 0.436 | 0.400 | 0.428 | 0.401 | 0.429 |
| Observations | 7366 | 7366 | 7366 | 7366 | 7366 | 7366 | 7366 | 7366 |

The sample comes from the 2012 and 2014 CFPS and includes children age between 6-144 months and their households. Children observed in year 2012 and 2014 are pooled together. "Age and gender" includes dummies of child age (in months) and dummy of child gender. Household covariates include household size, household location (rural or urban), father's and mother's education years and age, and the number of household members in the age categories 0-5, 6-15, 16-24, 25-49, a set of dummies indicating whether there is a women age over 50 and a man age over 50 within the household. "Older seniors" denotes the presence of males and females age over 70 within household separately. Robust standard errors are clustered at county level and presented in parenthesis. CFPS national sample weights in each year are used in the regressions.
\*\*\* $p < 0.01$ ; \*\* $p < 0.05$ ; \* $p < 0.1$

Specifications in columns 5-8 distinguish gender of pension recipient. Columns 5-6 included all baseline controls in column (3), and columns 7-8 follow column (4) in additionally controlling for presence of seniors age over 70. Age eligibility imposes essentially the same impact on male and female pension receipts, i.e., around 40 percentage points. Table A1 presents the results of household eligibility on NRPS pension receipt respectively for boy and girl subsamples.

### 5.2 Effects of NRPS on Child Weight

Table 3 reports the estimates on the impacts of NRPS on child BMI z scores. Columns 1-4 report estimates of equation (1) and columns 5-6 report estimates of equation (2). Panel A shows reduced-form estimates on pension age eligibility, and Panel B presents naive OLS estimates using pension receipt. Panel C presents 2SLS estimates using age eligibility as instruments for pension receipt. While naive OLS estimations report small and statistically insignificant effects, reduced-form estimations report sizable effects of NRPS on child weight. Specifically, average child BMI z scores in eligible households exceed those in ineligible households by 0.33-0.49 standard deviations (SD) across specifications in columns 1-4. The results are mostly statistically significant at 1% level. 2SLS estimates in Panel C report larger point estimates and standard errors. Impacts of receiving NRPS on child BMI z scores range from 0.74-1.09 SD. The baseline result in column 3 shows that NRPS increases child BMI by 0.91 SD for compliers.

**Table 3:** Effects of NRPS on Child BMI Z Score: Full Sample Results.

|  | (1) | (2) | (3) | (4) | (5) | (6) |
| --- | --- | --- | --- | --- | --- | --- |
| *Panel A: Reduced-form results* |  |  |  |  |  |  |
| NRPS eligibility | 0.395***<br>(0.126) | 0.334**<br>(0.150) | 0.412***<br>(0.138) | 0.496***<br>(0.156) |  |  |
| Male eligibility |  |  |  |  | 0.436***<br>(0.150) | 0.522***<br>(0.163) |
| Female eligibility |  |  |  |  | 0.038<br>(0.156) | 0.055<br>(0.169) |
| *Panel B: OLS results* |  |  |  |  |  |  |
| NRPS pensioner | 0.271*<br>(0.153) | 0.224<br>(0.155) | 0.193<br>(0.149) | 0.214<br>(0.153) |  |  |
| Male pensioner |  |  |  |  | -0.088<br>(0.167) | -0.082<br>(0.169) |
| Female pensioner |  |  |  |  | 0.258<br>(0.204) | 0.283<br>(0.206) |
| *Panel C: 2SLS results* |  |  |  |  |  |  |
| NRPS pensioner | 0.834***<br>(0.267) | 0.745**<br>(0.337) | 0.906***<br>(0.307) | 1.088***<br>(0.356) |  |  |
| Male pensioner |  |  |  |  | 1.092**<br>(0.427) | 1.301***<br>(0.447) |
| Female pensioner |  |  |  |  | -0.008<br>(0.404) | -0.016<br>(0.437) |
| Age and gender | Y | Y | Y | Y | Y | Y |
| Household covariates |  | Y | Y | Y | Y | Y |
| County FEs |  |  | Y | Y | Y | Y |
| Year FEs | Y | Y | Y | Y | Y | Y |
| Older seniors |  |  |  | Y |  | Y |
| Observations | 7366 | 7366 | 7366 | 7366 | 7366 | 7366 |

The sample comes from the 2012 and 2014 CFPS and includes children age between 6-144 months and their households. Children observed in year 2012 and 2014 are pooled together. "Age and gender" includes dummies of child age (in months) and dummy of child gender. Household covariates include household size, household location (rural or urban), father's and mother's education years and age, and the number of household members in the age categories 0-5, 6-15, 16-24, 25-49, a set of dummies indicating whether there is a women age over 50 and a man age over 50 within the household. "Older seniors" denotes the presence of males and females age over 70 within household separately. Robust standard errors are clustered at county level and presented in parenthesis. CFPS national sample weights in each year are used in the regressions.
\*\*\* $p < 0.01$ ; \*\* $p < 0.05$ ; \* $p < 0.1$

Columns 5-6 further distinguish the impacts of NRPS by gender of pensioners. Interestingly, similar to the reduced-form estimates, the effects of NRPS on child weight is almost entirely driven by grandfathers. With baseline controls, grandfathers receiving pension increases child BMI by 1.09 SD, about 20% larger than the baseline results without differentiating gender of pensioner. In contrast, the effects of receiving pension by grandmothers is close to zero and statistically insignificant. Additionally controlling for the presence of seniors over age 70 further magnifies the impact of male pensioners, with little change in the effect for female pensioners. The differential impacts of male and female pensioners are unlikely driven by differences in take-up rate, because Table 2 shows eligible males and females have the same propensity to enroll NRPS.

Our baseline effect size can be compared with those identified in previous studies. Duflo (2003)’s reduced-form estimates show social pension in South Africa increases girls’ weight-for-height z scores by 0.34, relative to 0.41 of BMI z scores in our baseline results (Table 3, Panel A, column (3)). Moreover, Duflo (2003) finds that the effect is driven by female pensioners, with a 2SLS estimate of 1.19 SD of weight-for-height. However, our study only shows male pensioners exert significant influence over child BMI, with a 2SLS estimate of 1.09 SD. Mu and de Brauw (2015) find rural parental migration in China increases child BMI by 0.11 SD, while Jo and Wang (2017) finds maternal full-time work raises urban Chinese child BMI by 1.11 SD. Therefore, our identified effect of NRPS receipt is larger than that of parental migration, but similar to the effects of maternal full-time work or grandparents receiving social pension in South Africa.

### 5.3 The Distributional Effects of NRPS on Child Weight

In addition to demonstrating that NRPS shapes overall child weight, we further evaluate its potential distributional effects on child weight. We replace the dependent variable in equations (1) and (2) with underweight, overweight and obesity, and report the linear probability model results in Table 4. All the columns report the 2SLS estimation results using baseline controls.

**Table 4:** Effects of NRPS on Child Underweight, Overweight and Obesity: Full Sample.

|  | Underweight |  | Overweight |  | Obesity |  |
| --- | --- | --- | --- | --- | --- | --- |
|  | (1) | (2) | (3) | (4) | (5) | (6) |
| NRPS pensioner | 0.002<br>(0.034) |  | 0.085**<br>(0.041) |  | 0.064**<br>(0.031) |  |
| Male pensioner |  | 0.023<br>(0.045) |  | 0.135**<br>(0.059) |  | 0.110***<br>(0.042) |
| Female pensioner |  | 0.005<br>(0.038) |  | -0.011<br>(0.058) |  | -0.014<br>(0.046) |
| Age and gender | Y | Y | Y | Y | Y | Y |
| Household covariates | Y | Y | Y | Y | Y | Y |
| County FEs | Y | Y | Y | Y | Y | Y |
| Year FEs | Y | Y | Y | Y | Y | Y |
| Observations | 7366 | 7366 | 7366 | 7366 | 7366 | 7366 |

This table presents the 2SLS estimates of NRPS on child weight outcomes. The sample comes from the 2012 and 2014 CFPS and includes children age between 6-144 months and their households. Children observed in year 2012 and 2014 are pooled together. "Age and gender" includes dummies of child age (in months) and dummy of child gender. Household covariates include household size, household location (rural or urban), father's and mother's education years and age, and the number of household members in the age categories 0-5, 6-15, 16-24, 25-49, a set of dummies indicating whether there is a women age over 50 and a man age over 50 within the household. Robust standard errors are clustered at county level and presented in parenthesis. CFPS national sample weights in each year are used in the regressions.
\*\*\* $p < 0.01$ ; \*\* $p < 0.05$ ; \* $p < 0.1$

We show that receiving pension has negligible impact on child underweight, while it increases the risks of child overweight and obesity. In particular, pension receipt increases rates of overweight and obesity by 27% (0.085/0.313) and 33% (0.064/0.195), respectively, and both are driven by male pensioners.

Overall, Tables 3 - 4 suggest that NRPS imposes significant impact on short-term nutritional outcomes among grandchildren. Grandfathers play a more important role than grandmothers do.

### 5.4 Effects of NRPS on Child Height

In this section, we evaluate how NRPS changes children’s longer term health outcomes as measured by height-for-age z scores. We replace the outcomes in equations (1)-(2) by child height-for-age z scores and report the results in Panel A, Table 5. All columns report 2SLS estimates with baseline controls.

**Table 5:** Effects of NRPS on Child Height.

|  | Full sample |  | Boys |  | Girls |  |
| --- | --- | --- | --- | --- | --- | --- |
|  | (1) | (2) | (3) | (4) | (5) | (6) |
| **Panel A: Height-for-age z-score** |  |  |  |  |  |  |
| NRPS pensioner | −0.308<br>(0.226) |  | −0.439<br>(0.346) |  | −0.410<br>(0.324) |  |
| Male pensioner |  | −0.205<br>(0.333) |  | −0.262<br>(0.492) |  | −0.089<br>(0.461) |
| Female pensioner |  | −0.135<br>(0.355) |  | −0.412<br>(0.528) |  | −0.167<br>(0.485) |
| **Panel B: Stunting** |  |  |  |  |  |  |
| NRPS pensioner | 0.032<br>(0.038) |  | 0.028<br>(0.061) |  | 0.041<br>(0.049) |  |
| Male pensioner |  | 0.039<br>(0.058) |  | 0.093<br>(0.077) |  | −0.050<br>(0.083) |
| Female pensioner |  | −0.024<br>(0.061) |  | −0.047<br>(0.096) |  | 0.050<br>(0.077) |
| Age and gender | Y | Y | Y | Y | Y | Y |
| Household covariates | Y | Y | Y | Y | Y | Y |
| County FEs | Y | Y | Y | Y | Y | Y |
| Year FEs | Y | Y | Y | Y | Y | Y |
| Observations | 7366 | 7366 | 3898 | 3898 | 3468 | 3468 |

This table presents the 2SLS estimates of NRPS on child height outcomes. The sample comes from the 2012 and 2014 CFPS and includes children age between 6-144 months and their households. Children observed in year 2012 and 2014 are pooled together. "Age and gender" includes dummies of child age (in months) and dummy of child gender. Household covariates include household size, household location (rural or urban), father's and mother's education years and age, and the number of household members in the age categories 0-5, 6-15, 16-24, 25-49, a set of dummies indicating whether there is a women age over 50 and a man age over 50 within the household. Robust standard errors are clustered at county level and presented in parenthesis. CFPS national sample weights in each year are used in the regressions.
\*\*\* $p < 0.01$ ; \*\* $p < 0.05$ ; \* $p < 0.1$

Columns 1-2 report full sample results respectively using equations (1) and (2). In both specifications the 2SLS estimates are imprecise to draw any statistical conclusion. Columns 3-4 and columns 5-6 further present the 2SLS estimates in boys and girls subsamples, respectively. Again, they are imprecisely estimated.

To explore the effects of NRPS on child stunting, an important measure of longterm impaired growth and development deficits, we repeat the exercise in Panel A upon replacing the outcome with a dummy variable indicating stunting. The results are reported in Panel B, Table 5. Again, none of the estimates are statistically significant. Results in Panel A and B, Table 5 both suggest NRPS may have little effect on child long-term nutritional status.

While the absence of an impact of pension income on height among grandchildren in rural China is at odds with Duflo (2003)’s finding that pension increases girls’ height-for-age z scores by 1.2 in South Africa, it is not surprising for at least two reasons. Firstly, social pension benefits only account for 10% of rural household income in China, too small to have any impact on child height in the short term. This is in stark contrast with South Africa in which pension benefits amount to more than twice the average rural income. Secondly, NRPS only completed its roll-out to all Chinese counties by the end of 2012, and by then a large proportion of households in each county had not enrolled in. Therefore, it can be too early to identify any long-term effect given the timing of CFPS 2012-2014.

### 5.5 Subsample Heterogeneity

We have presented so far that NRPS has substantial effects on child weight, but not on child height. The impacts are mostly driven by male pensioners. To further explore potential gender pattern in these identified effects, we divide the sample by child gender. In both subsamples we use baseline controls in estimations and differentiate gender of pensioners. The results are reported in Table 6.

**Table 6:**
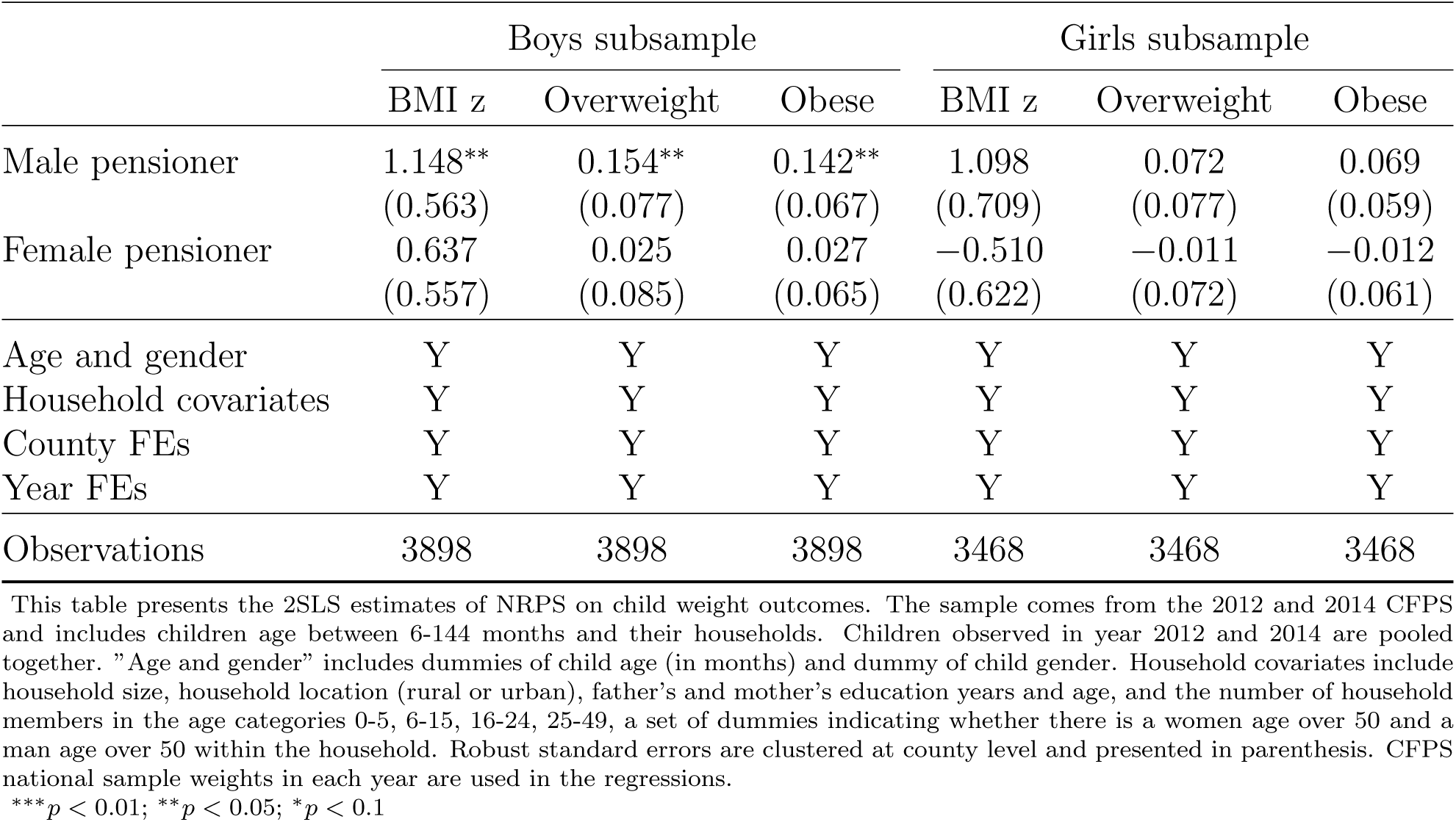
Effects of NRPS on Child Weight: Subsample Results by Child Gender.

|  | Boys subsample |  |  | Girls subsample |  |  |
| --- | --- | --- | --- | --- | --- | --- |
|  | BMI z | Overweight | Obese | BMI z | Overweight | Obese |
| Male pensioner | 1.148**<br>(0.563) | 0.154**<br>(0.077) | 0.142**<br>(0.067) | 1.098<br>(0.709) | 0.072<br>(0.077) | 0.069<br>(0.059) |
| Female pensioner | 0.637<br>(0.557) | 0.025<br>(0.085) | 0.027<br>(0.065) | -0.510<br>(0.622) | -0.011<br>(0.072) | -0.012<br>(0.061) |
| Age and gender | Y | Y | Y | Y | Y | Y |
| Household covariates | Y | Y | Y | Y | Y | Y |
| County FEs | Y | Y | Y | Y | Y | Y |
| Year FEs | Y | Y | Y | Y | Y | Y |
| Observations | 3898 | 3898 | 3898 | 3468 | 3468 | 3468 |

This table presents the 2SLS estimates of NRPS on child weight outcomes. The sample comes from the 2012 and 2014 CFPS and includes children age between 6-144 months and their households. Children observed in year 2012 and 2014 are pooled together. "Age and gender" includes dummies of child age (in months) and dummy of child gender. Household covariates include household size, household location (rural or urban), father's and mother's education years and age, and the number of household members in the age categories 0-5, 6-15, 16-24, 25-49, a set of dummies indicating whether there is a women age over 50 and a man age over 50 within the household. Robust standard errors are clustered at county level and presented in parenthesis. CFPS national sample weights in each year are used in the regressions.
\*\*\* $p < 0.01$ ; \*\* $p < 0.05$ ; \* $p < 0.1$

Interestingly, we find salient gender pattern in boys’ subsample with male pensioners. Benefits received by male pensioners is associated with boys’ larger BMI z score, rates of overweight and obesity. However, the effect of female pension receipt on boys’ weight are small and statistically indistinguishable. Meanwhile, in girls’ subsample, the estimates are imprecise, whether we examine the effects of benefits received by male or female pensioners. This is different from Duflo (2003)’s finding in South Africa that pension received by grandmothers promotes granddaughters’ weight.

### 5.6 Validity and Robustness

As our instrumental variable approach is essentially a Fuzzy Regression Discontinuity (RD) design around the pension age-eligibility threshold (60), scaled by the instrument, we show that our estimations strategy is valid, before testing robustness of our findings. First, our McCrary test (McCrary, 2008) in Figure A2 on the distribution of sampled population suggests no discontinuity around age 60 cut-off, which mitigates the concern over manipulation of age eligibility to receive NRPS. Second, observed pre-determined characteristics should have identical distributions on either side of the age cutoff. Figure A3 implements a placebo test of multi-generational co-residence before the introduction of NRPS. Reassuringly, results confirm continuity around age cut-off. We repeat this exercise for all pre-determined observables in our analysis. Results, upon request, suggest this is the case.

The 2SLS estimates compare compliers in treatment group with those in control group, which may not be generalized to the group of non-compliers. To evaluate the average effect between compliers and non-compliers, intent-to-treat (ITT) estimates are presented in Table A2 that compare between children in eligible households with those in ineligible households before and after NRPS expansion.^8^ CFPS collected first wave of data in 2010, when NRPS coverage was very low.^9^ Our DD estimates of the effects of NPRS on child weight are reported in Table A2. The results, while attenuated by non-compliers, are largely consistent with our findings in Table 3.

Finally, The presence of NRPS pensioner in a household may be endogenous. In particular, household composition, i.e. living arrangement of grandparents with their grandchildren, may also change with pension roll-out, which undermines the exogeneity of the presence of NRPS eligible household members. In other words, endogenous household composition could create a correlation between unobserved household characteristics and the presence of an eligible member, which may invalidate our proposed identification strategy of pension receipt on nutritional status of grandchildren. Following Duflo (2003)’s proposed solution, we re-construct household pension age eligibility that includes all extended family members, regardless of their co-residence status. Results, shown in Table A3, are qualitatively similar to our main estimates, suggesting the validity of our 2SLS estimations. The smaller marginal effect of pension receipt may reflect the fact that grandparents living away from grandchildren may impose less influence over their nutritious status.

## 6 Mechanisms and Other Outcomes

In a household decision-making framework, children are considered public good. Pension benefits to grandparents may shape health outcomes among grandchildren in two main channels, goods and time allocated to grandchildren. In this section, we examine these plausible mechanisms.

### 6.1 Income Effect

While we are unable to directly test how NRPS changes child consumption as CFPS does not survey on individual consumption or specifically nutritional intake, we first test whether NRPS discontinuously increases sources of household income around age 60. As food consumption is considered a normal good, income expansion to grandparents has the potential to increase food consumption allocated to grandchildren.

We extract the adult sample from CFPS waves 2012-2014, centering at 60 and coding their age in 0.5 years. We match the adults observations age 50-70 with their household income data. Adults exactly age 60 are dropped. In total 10254 adults are in our sample. Figure 3a displays the NRPS take-up rate by age. There is an immediate and discontinuous increase in the take-up rate at age 60. We use a RD design and local linear regression to estimate the impacts of NRPS (see Imbens and Lemieux (2008)). Triangular kernel is used in the estimations. Adult and household characteristics in our baseline model are controlled for in all RD estimations here.

RD estimates show that NRPS increases annual household income per capita by more than 750 CNY, which accounts for above 10 % of our sample average income and above 15% of the median income in our sample, which is similar to Chinese official statistics that NRPS pension income accounts for about 10% of rural household income per capita in 2012.^10^ To confirm the sources of rising income, we further decompose household income into five categories: public transfer, wage income, capital income, household business income and other income.^11^ The results are shown in Figure A4a - A4e. NRPS increases public transfer to households by 219 CNY on average, and it also increases household capital income by 24 CNY. The rising capital income may due to the increase in household savings after receiving NRPS. The impacts of NRPS on other sources of income are statistically insignificant. In addtion to household income, Figure A4f displays the impacts of NRPS on individual income.^12^ Our estimate suggests that NRPS increases individual income by more than 415 CNY, an impact smaller but consistent with our findings on household income.

### 6.2 Time allocation

Time allocated to child care may also be affected by NRPS. For instance, grandparents may reduce labor supply after receiving pension, and instead spend more time with grandchildren, which frees up parents’ time in housework and may increase their labor supply. If this is the case, we may not necessarily observe substantial increase in household total wage income, other than increase in grandparents’ time allocated to grandchildren and their nutritional outcomes. While household time allocation is not directly observed in CFPS, other indirect measures, i.e., adult child migration and main caregivers for grandchildren, may indicate grandparents’ time allocation to grandchildren.

Figure 4a displays the RD estimates on NRPS receipt and adult child migration. Adult child migration is defined based on adult children of older adults leaving the home county for 3 months and more in a year.

**Figure 4:**
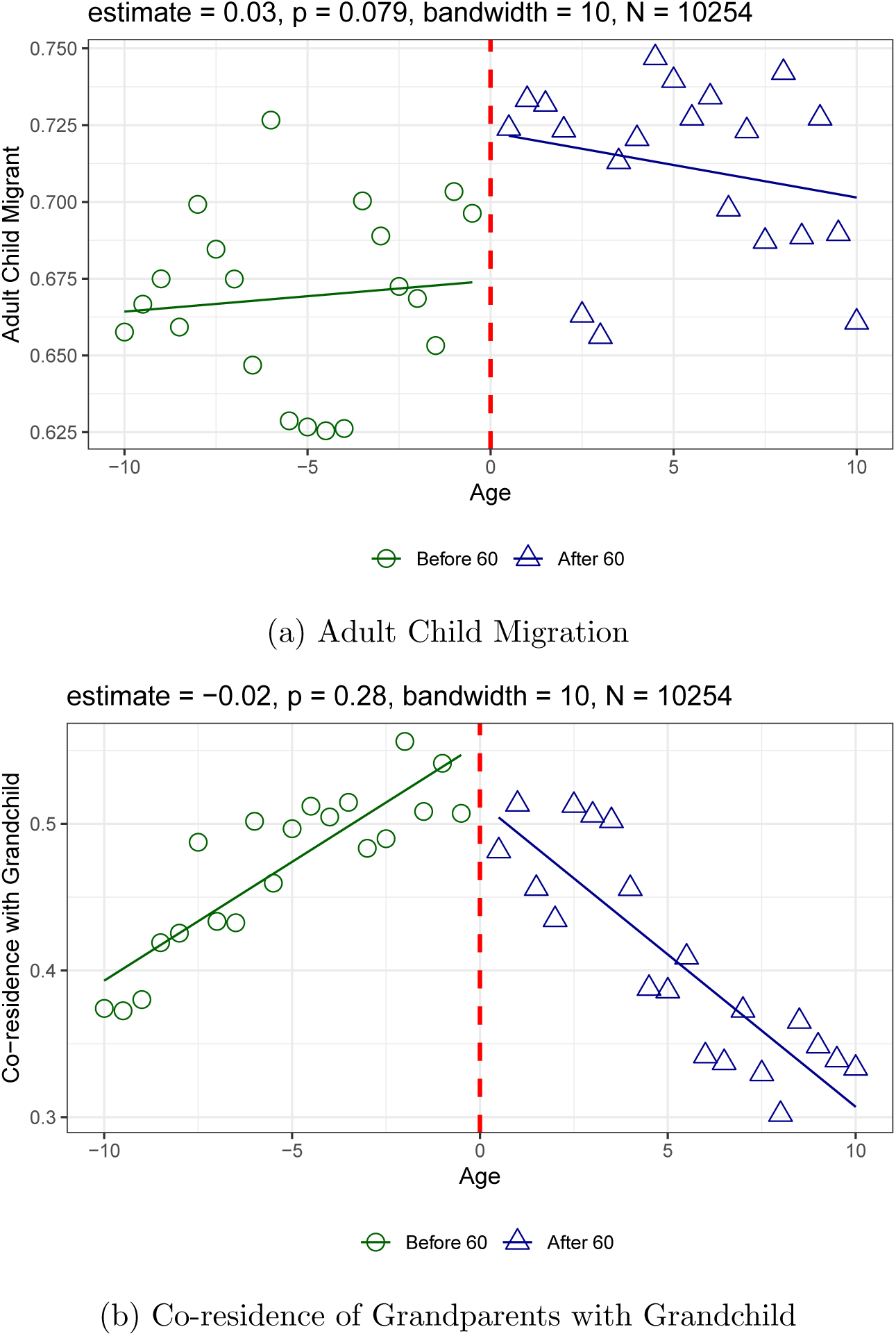
Effects of NRPS on Adult Child Migration and Multi-generational Co-residence. *Note*: The data comes from CFPS wave 2012 and 2014. Ages are centered at 60. Panel (a) shows the change of the grandparents’ co-residence rate with child under 12 by grandparent’s age (in 0.5 years). Panel (b) shows the change of the grandparents’ adult child migration rate by grandparents’ age (in 0.5 years). The solid lines show the fitted lines before and after 60 years old. The RD estimation results are displayed in each panel using adult observations.

We show a 3 percentage point increase in adult child migration following pension receipt. The result is consistent with Eggleston, Sun and Zhan (2018)’s study, where they find a larger impact of NRPS on adult child migration, especially in less developed areas in China. Our identified small impact on adult child migration could be explained by CFPS inclusion of rural counties in both more developed and less developed areas. Our finding implies that the burden of child care may shift from parents to grandparents after parental migration. However, this modest effect is unlikely to drive the identified substantial effect on child weight and gender differences.

CFPS surveys include a question regarding main caregivers in daytime and at night for children under 12. In Table 7, we re-estimate equation (2) by replacing the outcome with whether the child is mainly taken care of by grandparents. The results are largely insignificant in both boys and girls subsamples, with the only exception being that grandmothers’ pension marginally increases their serving as the main caregivers for granddaughters in daytime. Overall, our empirical evidence suggests that the shift of care-giving responsibility from parents to grandparents may not be large enough to explain our main findings on gendered pattern in nutritional outcomes among grandchildren.

**Table 7:** Effects of NRPS on Grandparents being the Primary Child Caregivers.

|  | Full sample |  | Boys |  | Girls |  |
| --- | --- | --- | --- | --- | --- | --- |
|  | Daytime | Night | Daytime | Night | Daytime | Night |
| Male pensioner | 0.044<br>(0.068) | -0.039<br>(0.065) | 0.090<br>(0.088) | -0.015<br>(0.097) | 0.013<br>(0.091) | -0.006<br>(0.082) |
| Female pensioner | 0.031<br>(0.068) | -0.053<br>(0.062) | -0.048<br>(0.076) | -0.104<br>(0.081) | 0.184*<br>(0.101) | 0.016<br>(0.098) |
| Age and gender | Y | Y | Y | Y | Y | Y |
| Household covariates | Y | Y | Y | Y | Y | Y |
| County FEs | Y | Y | Y | Y | Y | Y |
| Year FEs | Y | Y | Y | Y | Y | Y |
| Observations | 7365 | 7365 | 3897 | 3897 | 3468 | 3468 |

This table presents the 2SLS estimates of NRPS on child care-giving arrangements. Daytime child caregiver is defined by CFPS question "who is usually the main caregiver of the child during the daytime?" Night child caregiver is defined by CFPS question "who is usually the main caregiver of the child during the night?" The sample comes from the 2012 and 2014 CFPS and includes children age between 6-144 months and their households. Children observed in year 2012 and 2014 are pooled together. "Age and gender" includes dummies of child age (in months) and dummy of child gender. Household covariates include household size, household location (rural or urban), father's and mother's education years and age, and the number of household members in the age categories 0-5, 6-15, 16-24, 25-49, a set of dummies indicating whether there is a women age over 50 and a man age over 50 within the household. Robust standard errors are clustered at county level and presented in parenthesis. CFPS national sample weights in each year are used in the regressions.
\*\*\* $p < 0.01$ ; \*\* $p < 0.05$ ; \* $p < 0.1$

### 6.3 Son Preference

Our findings of the link between grandfathers’ pension receipt and grandsons’ health outcomes may indicate the prevailing son preference in rural China, and grandfathers may possess stronger norms of son preference than grandmothers in a traditional patrilineal society. A potential way to help test son preference is to further differentiate by gender of the intermediate generation. We predict that pension receipt by father’s father may have stronger impacts on grandson’s weight than that by mother’s father, because the family name is carried on only through males. Table 8 reports the reduced-form OLS estimates by differentiating the gender of intermediate generation. As expected, results show that the effect through father’s father on boys weight are relatively more salient. The effects of pension receipt by father’s father on boys over-weight/obesity are statistically significant at 10% level. The effects through mother’s father are also positive but imprecisely estimated.

**Table 8:** Effects of NRPS on Child Weight by Gender of Intermediate Generation.

|  | Full sample |  |  | Boys |  |  | Girls |  |  |
| --- | --- | --- | --- | --- | --- | --- | --- | --- | --- |
|  | BMI z | Overweight | Obese | BMI z | Overweight | Obese | BMI z | Overweight | Obese |
| Farther's father eligible | 0.501**<br>(0.218) | 0.054**<br>(0.026) | 0.052**<br>(0.022) | 0.453<br>(0.330) | 0.066*<br>(0.033) | 0.057*<br>(0.034) | 0.441<br>(0.281) | 0.011<br>(0.035) | 0.041<br>(0.029) |
| Farther's mother eligible | 0.051<br>(0.180) | 0.005<br>(0.027) | 0.004<br>(0.020) | 0.365<br>(0.317) | 0.026<br>(0.043) | 0.030<br>(0.033) | -0.060<br>(0.227) | 0.006<br>(0.033) | -0.004<br>(0.025) |
| Mother's father eligible | 0.501<br>(0.487) | 0.039<br>(0.067) | 0.025<br>(0.071) | 0.826<br>(0.623) | 0.057<br>(0.087) | 0.096<br>(0.097) | 0.406<br>(0.625) | 0.041<br>(0.108) | 0.031<br>(0.086) |
| Mother's mother eligible | 0.417<br>(0.683) | 0.114<br>(0.083) | 0.030<br>(0.089) | 0.595<br>(0.983) | 0.045<br>(0.124) | 0.079<br>(0.130) | 0.685<br>(0.792) | 0.172<br>(0.119) | -0.019<br>(0.102) |
| Age and gender | Y | Y | Y | Y | Y | Y | Y | Y | Y |
| Household covariates | Y | Y | Y | Y | Y | Y | Y | Y | Y |
| County FEs | Y | Y | Y | Y | Y | Y | Y | Y | Y |
| Year FEs | Y | Y | Y | Y | Y | Y | Y | Y | Y |
| R<sup>2</sup> | 0.218 | 0.188 | 0.186 | 0.227 | 0.213 | 0.212 | 0.297 | 0.268 | 0.267 |
| Observations | 7366 | 7366 | 7366 | 3898 | 3898 | 3898 | 3468 | 3468 | 3468 |

This table presents the OLS estimates of NRPS eligibility on child weight outcomes, by differentiating father's parents and mother's parents. The sample comes from the 2012 and 2014 CFPS and includes children age between 6-144 months and their households. Children observed in year 2012 and 2014 are pooled together. "Age and gender" includes dummies of child age (in months) and dummy of child gender. Household covariates include household size, household location (rural or urban), father's and mother's education years and age, and the number of household members in the age categories 0-5, 6-15, 16-24, 25-49, a set of dummies indicating whether there is a woman age over 50 and a man age over 50 within the household. Robust standard errors are clustered at county level and presented in parenthesis. CFPS national sample weights in each year are used in the regressions.
\*\*\* $p < 0.01$ ; \*\* $p < 0.05$ ; \* $p < 0.1$

### 6.4 Co-residence Behavior

The mechanisms so far largely focus on co-residing families, as in our main results we have been using co-residing household members’ pension eligibility as the treatment. The co-residence behavior of grandparents with grandchildren could re-enhance these mechanisms through an “extensive margin”. In CFPS 2012-2014, around 45% of elderly at age 60 co-reside with grandchildren under 12, a significant jump from around 37% in 2010.

We next explore the potential impact of NRPS on multi-generational co-residence decisions and show the results in Figure 4b. Multi-generational co-residence is defined as grandparents living with children under 12. The RD estimates using CFPS 2012-2014 show that the impact of NRPS on co-residence of grandparents with grand-children is small in size and statistically insignificant. We replicate the estimation in a sample of non-NRPS counties in CFPS 2010 and find similar results (see Figure A3). The results reassure us that the time trend of multi-generational co-residence rate prior to NRPS is not driven by its anticipated roll-out, if any.

## 7 Conclusion

In this paper, we present novel evidence on the multi-generational health effects of the largest social pension policy in the world, i.e., China’s NRPS. We take advantage of the discontinuity in age eligibility for pension receipt, and find robust results that NRPS impose substantial effects on grandchildren’s short-term nutritional outcomes. The effect seems driven by grandfathers on grandsons.

While we discuss some of the potential mechanisms that the effect can be plausibly explained by NRPS increasing child food consumption, exacerbated via son preference and modest increase in grandparents’ time allocation to child care, we leave the definitive explanations unexplored. For instance, our findings could not distinguish if the differential effects by gender of grandchildren emerge from grandparents’ preference or from perceived differences in the returns to inputs. If the former dominates, future work needs to understand the exact cause(s) of differential preferences.

Our findings are not as promising as it seems, as we show NRPS increases children’s chance of overweight and obesity, but does not reduces their underweight rate. In the meantime, NRPS does not seem to improve children’s long-term nutritional outcomes. Therefore, our findings lend support to the ever-increasing concern over double burden of under-and over-nutrition, especially in less developed areas where grand-parents spend more time on child care with very limited knowledge about healthy diet and physical activities for children. In order to improve the heath outcomes of rural children, relying on public transfer alone without addressing the key issue of family decision-making on child rearing may not be sufficient.

## Data Availability

All data produced in the present study are available upon reasonable request to the authors.

https://www.isss.pku.edu.cn/cfps/en/index.htm?CSRFT=Q9BL-UGK3-26PK-N7FS-QJW9-V1DT-5JX5-U7RI

## Appendix Figures and Tables

**Figure A1:**
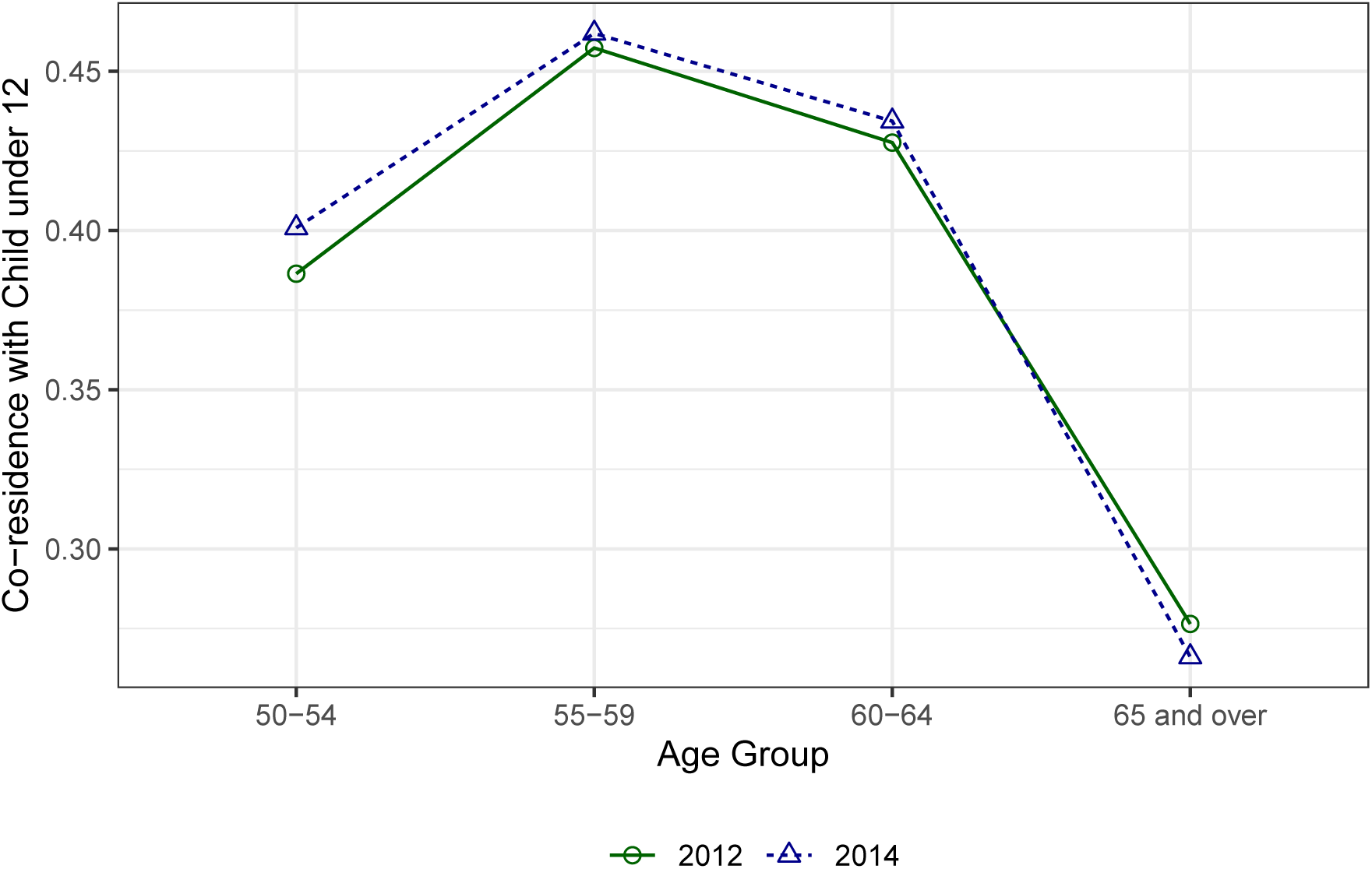
Share of Grandparents Co-residing with Grandchildren under Age 12. *Source:* Authors’ tabulations of China Family Panel Studies waves 2012 and 2014.

**Figure A2:**
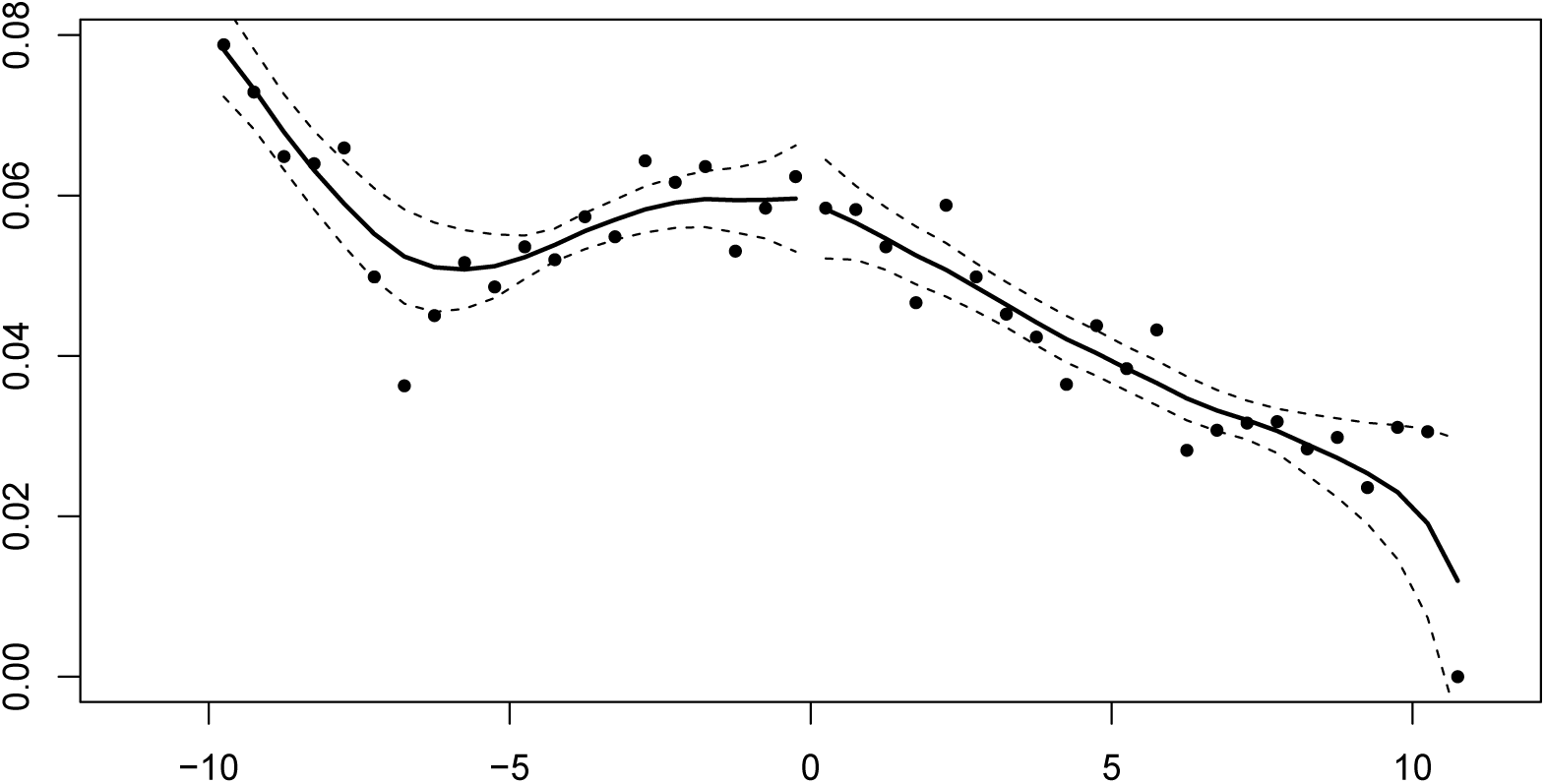
McCrary Sorting Test: P-value = 0.93. *Note*: The sample comes from CFPS wave 2012 and 2014. Each dot represents the data frequency at each age (in 0.5 years). Ages are centered at 60 years old. The solid line displays the local linear regression of frequencies on age, before and after 60 years old. The null hypothesis is that the distribution of sample frequency at the cutoff age 60 is continuous.

**Figure A3:**
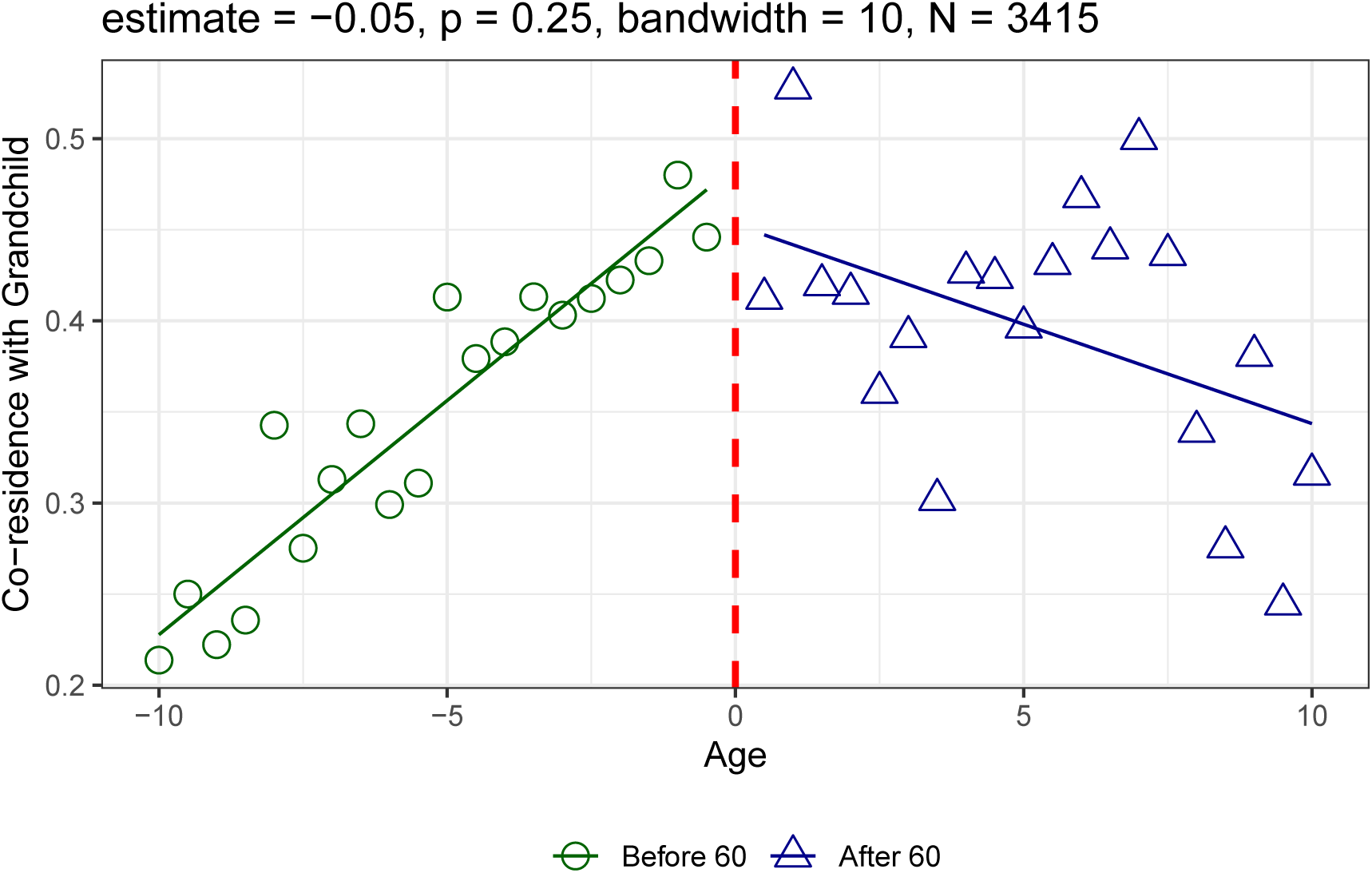
Testing the Placebo Effect of NRPS on Multi-generational Co-residence Prior to NRPS roll-out. *Note*: The data comes from CFPS wave 2010, and only includes households in counties prior to the NRPS roll-out. Ages are centered at 60. The solid lines show the fitted lines before and after 60 years old. The RD estimation results are displayed in each panel using adult observations.

**Figure A4:**
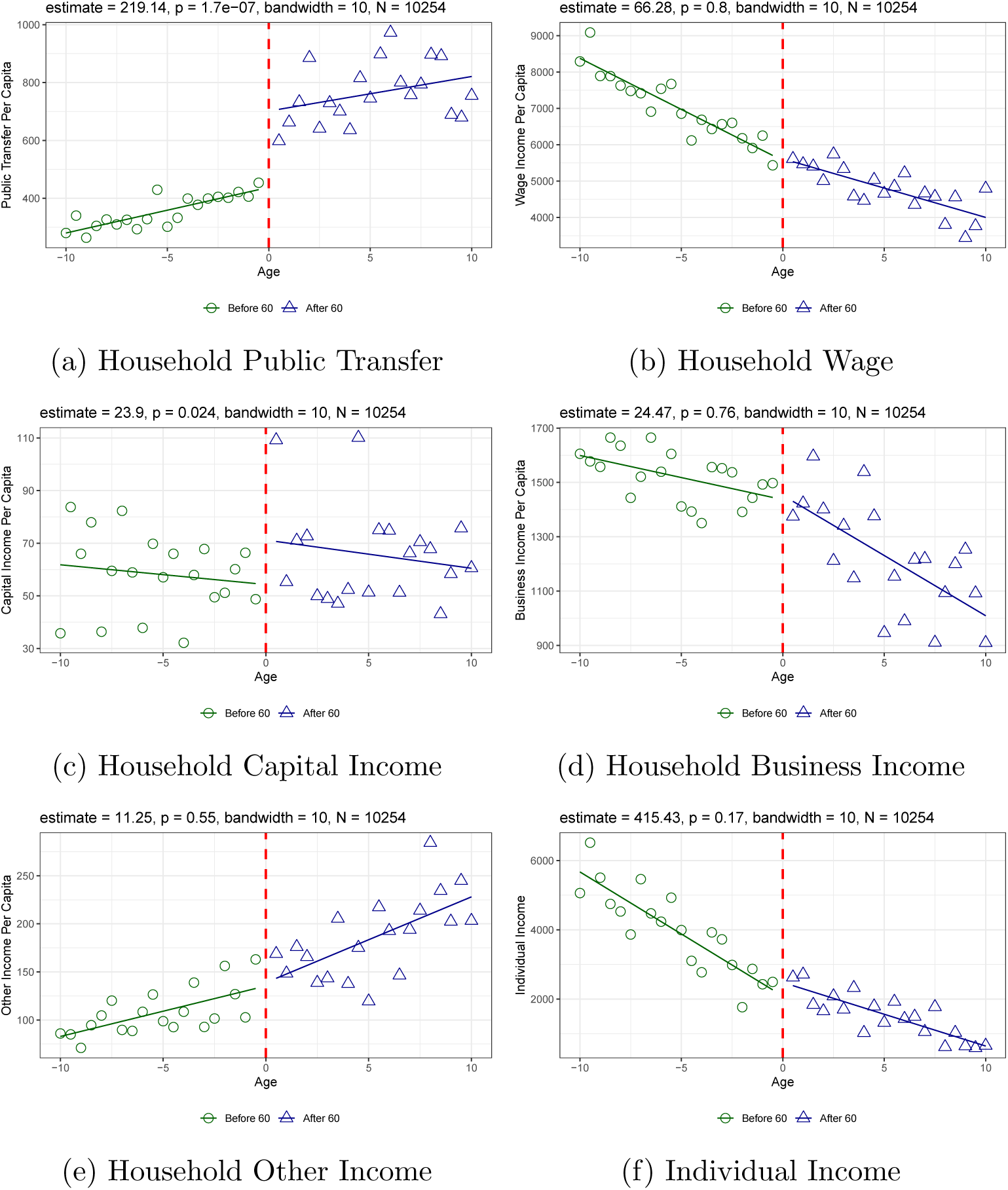
Effects of NRPS on Household Income: by Income Sources. *Note*: The data comes from CFPS wave 2012 and 2014. Ages are centered at 60. Panels (a)-(e) displays the average household income per capita by income sources at ages (in 0.5 years) 50-70. Household income per capita is divided by five incomes sources. Household public transfer includes all pension, subsidies and compensations as well as income from public donation. Household wage includes all the wage from each household member. Household captial income includes all gains from financial investment and rental income from real estate properties, land, and machineries. Household business income includes all net income from family agricultural work (including in-kind income), and net profit from family-owned businesses. Household other income includes all monetary support from friends and relatives. Panel (f) displays the average adult individual income at ages (in half years) 50-70. For each adult, individual income consists of income from internships, full-time work, pension, fellowship, and assistantship. The solid lines show the fitted lines before and after 60 years old. The RD estimation results are displayed in each panel using adult observations.

**Table A1:** Effects of Age Eligibility on Pension Receipt: Sub-sample Results by Gender of Children.

|  | Either gender |  | Male |  | Female |  |
| --- | --- | --- | --- | --- | --- | --- |
|  | Boys | Girls | Boys | Girls | Boys | Girls |
|  | (1) | (2) | (3) | (4) | (5) | (6) |
| NRPS eligibility | 0.448***<br>(0.035) | 0.465***<br>(0.030) |  |  |  |  |
| Male eligibility |  |  | 0.386***<br>(0.038) | 0.417***<br>(0.033) | 0.046**<br>(0.023) | 0.064**<br>(0.024) |
| Female eligibility |  |  | 0.025<br>(0.023) | 0.049**<br>(0.024) | 0.413***<br>(0.031) | 0.396***<br>(0.029) |
| Age and gender | Y | Y | Y | Y | Y | Y |
| Household covariates | Y | Y | Y | Y | Y | Y |
| County FEs | Y | Y | Y | Y | Y | Y |
| Year FEs | Y | Y | Y | Y | Y | Y |
| F statistics | 163.84 | 240.25 | 103.18 | 159.68 | 177.49 | 186.46 |
| R<sup>2</sup> | 0.459 | 0.466 | 0.421 | 0.452 | 0.464 | 0.447 |
| Observations | 3898 | 3468 | 3898 | 3468 | 3898 | 3468 |

This table presents the OLS estimates of NRPS eligibility on pension receipt. The sample comes from the 2012 and 2014 CFPS and includes children age between 6-144 months and their households. Children observed in year 2012 and 2014 are pooled together. "Age and gender" includes dummies of child age (in months) and dummy of child gender. Household covariates include household size, household location (rural or urban), father's and mother's education years and age, and the number of household members in the age categories 0-5, 6-15, 16-24, 25-49, a set of dummies indicating whether there is a women age over 50 and a man age over 50 within the household. Robust standard errors are clustered at county level and presented in parenthesis. CFPS national sample weights in each year are used in the regressions.
\*\*\* $p < 0.01$ ; \*\* $p < 0.05$ ; \* $p < 0.1$

**Table A2:** Effects of NRPS on Child BMI Z Score: Difference-in-Difference Estimation.

|  | Full | Full | Boys | Girls |
| --- | --- | --- | --- | --- |
| Eligible household × After county rollout | 0.456**<br>(0.228) | | | |
| Eligible household | -0.050<br>(0.232) |  |  |  |
| Eligible male members × After county rollout | | 0.338<br>(0.282) | 0.632*<br>(0.359) | 0.023<br>(0.375) |
| Eligible female members × After county rollout | | 0.238<br>(0.329) | -0.045<br>(0.520) | 0.420<br>(0.356) |
| Eligible male members |  | -0.072<br>(0.278) | -0.330<br>(0.343) | 0.157<br>(0.400) |
| Eligible female members |  | -0.046<br>(0.309) | 0.443<br>(0.491) | -0.402<br>(0.363) |
| Age and gender | Y | Y | Y | Y |
| Household covariates | Y | Y | Y | Y |
| County FEs | Y | Y | Y | Y |
| Year FEs | Y | Y | Y | Y |
| R<sup>2</sup> | 0.200 | 0.200 | 0.199 | 0.268 |
| Observations | 11051 | 11051 | 5892 | 5159 |

The sample comes from the CFPS 2010, 2012 and 2014, and it includes children age between 6-144 months and their households. Children observed in all years are pooled together. "After country rollout" indicate whether the county a household located in has rolled-out NRPS in the survey year or not. The county NRPS rollout year is constructed using the 5th percentile of household NRPS participation years within the county. "Age and gender" includes dummies of child age (in months) and dummy of child gender. Household covariates include household size, household location (rural or urban), father's and mother's education years and age, and the number of household members in the age categories 0-5, 6-15, 16-24, 25-49, a set of dummies indicating whether there is a women age over 50 and a man age over 50 within the household. Robust standard errors are clustered at county level and presented in parenthesis. CFPS national sample weights in each year are used in the regressions.
\*\*\* $p < 0.01$ ; \*\* $p < 0.05$ ; \* $p < 0.1$

**Table A3:** Effects of NRPS on BMI Z Scores, Regardless of Co-residence Status.

|  | Full | Full | Boys | Girls |
| --- | --- | --- | --- | --- |
| NRPS pensioner | 0.793**<br>(0.327) |  |  |  |
| Male pensioner |  | 0.926*<br>(0.473) | 0.962*<br>(0.570) | 1.002<br>(0.852) |
| Female pensioner |  | 0.090<br>(0.436) | 0.898<br>(0.572) | -0.545<br>(0.717) |
| Age and gender | Y | Y | Y | Y |
| Household covariates | Y | Y | Y | Y |
| County FEs | Y | Y | Y | Y |
| Year FEs | Y | Y | Y | Y |
| R<sup>2</sup> | 0.211 | 0.208 | 0.212 | 0.284 |
| Observations | 7366 | 7366 | 3898 | 3468 |

This table presents the 2SLS estimates of NRPS on child weight outcomes. The sample comes from the CFPS 2012 and 2014, and it includes children age between 6-144 months and their households. Children observed in all years are pooled together. Household eligibility is constructed by whether children's extended family members in the sample, regardless of their co-residence status, are eligible to receive NRPS. "Age and gender" includes dummies of child age (in months) and dummy of child gender. Household covariates include household size, household location (rural or urban), father's and mother's education years and age, and the number of household members in the age categories 0-5, 6-15, 16-24, 25-49, a set of dummies indicating whether there is a women age over 50 and a man age over 50 within the household. Robust standard errors are clustered at county level and presented in parenthesis. CFPS national sample weights in each year are used in the regressions.
\*\*\* $p < 0.01$ ; \*\* $p < 0.05$ ; \* $p < 0.1$

## Footnotes

1 Hukou system is an official household registration system that divides residents mainly into rural and urban types.

2 In our sample, 1444 out of 7366 children (or 19.6% of the sample) live with NRPS recipient, among whom 553 children (38.3%) report grandparents as their main daycare givers, and 478 children (33.1%) report grandparents as their main night-care givers.

3 At the beginning of NRPS roll-out, a family binding policy was in place requiring pension recipients to also enroll their eligible adult children to contribute premium to their personal pension account. Nonetheless, since even the minimum annual non-contributory benefits (660 yuan) was much larger than the minimum annual premium (100 yuan) paid by children, the two generations still had incentive to enroll. The binding policy was later removed.

4 Rural Migrant Monitoring Report, National Bureau of Statistics: http://www.stats.gov.cn/tjsj/zxfb/202104/t20210430_1816933.html

5 The lack of equal opportunities for rural migrants to access public services, such as child schooling, unemployment supports, health care and retirement security, strongly discourages them from migrating with family (Song, 2014; Au and Henderson, 2006; Meng, 2012). In 2015, over 40.5 million rural children under 17 live in their original domicile without either or both parents due to parental migration. The number of left behind children comes from United Nations Children’s Fund (UNICEF) Annual Report 2015 China. The number for rural compulsory school children left behind in 2017 is 15.5 millions, according to our tabulation of the educational statistics released by Ministry of Education. The original data can be seen for primary and secondary school children left behind http://www.moe.gov.cn/jyb_sjzl/moe_560/jytjsj_2017.

6 This study includes all children under 12. We measure children’s BMI z score and height-for-age z score based on Child Growth Standards (0-5 years) and Growth Reference (5-19 years) developed by the World Health Organization (WHO). We do not use weight-for-height z score because WHO only has weight-for-height growth standard for children under 5.

7 NRPS eligibility depends on the rural hukou type, rather than residence location.

8 CFPS longitudinal survey covers the gradual roll-out of NRPS, which enables us to leverage policy dynamics in a standard Difference-in-Difference (DD) analysis. In contrast to the cross-sectional setting in Duflo (2003)’s DD evaluation of social pension in South Africa that rely on individual-level NRPS take-up decisions, our DD analysis makes use of county-level roll-out timing. Yang and Bazan Ruiz (2021) find while the variation of county roll-out timing is partially driven by local economic and political forces, most of the variation remains unexplained, which supports the exogeneity of using county roll-out timing.

9 In comparison to our main 2SLS estimations, our DD analysis enables further inclusion of CFPS 2010 survey, prior to NRPS roll-out in a majority of CFPS counties.

10 Household income per capita is calculated by household gross income divided by number of household members.

11 Household public transfer includes all pension, subsidies and compensations as well as income from public donation. Household wage income includes all wages from household members. House-hold capital income includes all gains from financial investment and rental income from real estate properties, land, and machineries. Household business income includes all net income from family agricultural work (including in-kind income), and net profit from family-owned businesses. House-hold other income includes all monetary support from friends and relatives.

12 Individual income consists of income from internships, full-time work, pension, fellowship, and assistantship.

## Notes

‡ This work was supported by a number of NIH grants (K01AG053408; P30AG021342; P30AG066508); James Tobin Research Fund at Yale Economics Department; and Yale Macmillan Center Faculty Research Award. The funders had no role in the study design; data collection, analysis, or interpretation; in the writing of the report; or in the decision to submit the article for publication. The authors acknowledge the Institute of Social Science Survey at Peking University for providing us with the CFPS data, which are available online at CFPS official website. The authors also acknowledge helpful comments by participants and discussants at the various conferences, seminars and workshops. The authors have no conflict of interest. The study was approved by the Institutional Review Board (IRB) at Peking University (Approval No: IRB00001052-14010). All participants gave informed consent in accordance with policies of the IRB at Peking University.

### Competing Interest Statement

The authors have declared no competing interest.

### Funding Statement

This work was supported by a number of NIH grants (K01AG053408; P30AG021342; P30AG066508); James Tobin Research Fund at Yale Economics Department; and Yale Macmillan Center Faculty Research Award.

### Author Declarations

The source data - CFPS - are available to the public before the initiation of the study. The Institute of Social Science Survey at Peking University provides us with the CFPS data, which are available online at CFPS official website.

## References

Aizer, Anna, Shari Eli, Joseph Ferrie, and Adriana Lleras-Muney. 2016. “The Long Term Impact of Cash Transfers to Poor Families.” American Econmic Review, 106(4): 935–971.

Au, Chun-Chung, and J Vernon Henderson. 2006. “Are Chinese Cities too Small?.” Review of Economic Studies, 73(3): 549–576.

Case, Anne. 2001. “Does Money Protect Health Status? Evidence from South African Pensions.” NBER working paper, No. 8495.

Case, Anne, and Angus Deaton. 1998. “Large Cash Transfers to the Elderly in South Africa.” The Economic Journal, 108(450): 1330–1361.

Cawley, John. 2015. “An Economy of Scales: A Selective Review of Obesity’s Economic Causes, Consequences, and Solutions.” Journal of Health Economics, 43 244–268.

Chen, Xi, Karen Eggleston, and Ang Sun. 2017. “The impact of social pensions on intergenerational relationships: Comparative evidence from China.” Journal of the Economics of Ageing, 12 225–235.

Chen, Xi, Lipeng Hu, and Jody L. Sindelar. 2020. “Leaving money on the table? Suboptimal enrollment in the new social pension program in China.” The Journal of the Economics of Ageing, 15, p. 100233.

Chen, Xi, Tianyu Wang, and Susan H Busch. 2019. “Does Money Relieve Depression? Evidence from Social Pension Expansions in China.” Social Science & Medicine, 220 411–420.

Cheng, Lingguo, Hong Liu, Ye Zhang, and Zhong Zhao. 2018a. “The Health Implications of Social Pensions: Evidence from China’s New Rural Pension Scheme.” Journal of Comparative Economics, 46(1): 20–34.

Cheng, Lingguo, Hong Liu, Ye Zhang, and Zhong Zhao. 2018b. “The Heterogeneous Impact of Pension Income on Elderly Living Arrangements: Evidence from China’s New Rural Pension Scheme.” Journal of Population Economics, 31(1): 155–192.

Dizon-Ross, Rebecca, and Seema Jayachandran. 2022. “Dads and Daughters: Disentangling Altruism and Investment Motives for Spending on Children.” NBER Working Paper, No. 29912.

Duflo, Esther. 2003. “Grandmothers and Granddaughters: Old-Age Pensions and Intrahousehold Allocation in South Africa.” World Bank Economic Review, 17(1): 1–25.

Duflo, Esther. 2012. “Women Empowerment and Economic Development.” Journal of Economic Literature, 50(4): 1051–1079.

Duflo, Esther, and Christopher Udry. 2004. “Intrahousehold Resource Allocation in Côte D’Ivoire: Social Norms, Separate Accounts and Consumption Choices.” NBER working paper, No. 10498.

Eggleston, Karen, Ang Sun, and Zhaoguo Zhan. 2018. “The impact of rural pensions in China on labor migration.” World Bank Economic Review, 32(1): 64–84.

He, Qinying, Xun Li, and Rui Wang. 2018. “Childhood obesity in China: Does grandparents’ coresidence matter?.” Economics & Human Biology, 29 56–63.

Huang, Wei, and Chuanchuan Zhang. 2021. “The Power of Social Pensions: Evidence from China’s New Rural Pension Scheme.” American Economic Journal: Applied Economics, 13(2): 179–205.

Imbens, Guido W., and Thomas Lemieux. 2008. “Regression discontinuity designs: A guide to practice.” Journal of Econometrics, 142(2): 615–635.

Jensen, Robert T. 2004. “Do Private Transfers ‘Displace’ the Benefits of Public Transfers? Evidence from South Africa.” Journal of Public Economics, 88(1-2): 89–112.

Jo, Young, and Qing Wang. 2017. “The impact of maternal employment on children’s adiposity: Evidence from China’s labor policy reform.” Health Economics, 26(12): e236–e255.

Lundberg, S. 2005. “Sons, Daughters, and Parental Behaviour.” Oxford Review of Economic Policy, 21 340–356.

Maitra, Pushkar, and Ranjan Ray. 2003. “The Effect of Transfers on Household Expenditure Patterns and Poverty in South Africa.” Journal of Development Economics, 71(1): 23–49.

McCrary, Justin. 2008. “Manipulation of the running variable in the regression discontinuity design: A density test.” Journal of Econometrics, 142(2): 698–714.

Meng, Xin. 2012. “Labor Market Outcomes and Reforms in China.” Journal of Economic Perspectives, 26(4): 75–102.

Mu, Ren, and Alan de Brauw. 2015. “Migration and Young Child Nutrition: Evidence from Rural China.” Journal of Population Economics, 28(3): 631–657.

Nikolov, Plamen, and Alan Adelman. 2019. “Do private household transfers to the elderly respond to public pension benefits? Evidence from rural China.” The Journal of the Economics of Ageing, 14, p. 100204.

Ning, Manxiu, Jinquan Gong, Xuhui Zheng, and Jun Zhuang. 2016. “Does New Rural Pension Scheme Decrease Elderly Labor Supply? Evidence from CHARLS.” China Economic Review, 41 315–330.

Piernas, C., D. Wang, S. Du, B. Zhang, Z. Wang, C. Su, and B. M. Popkin. 2015. “The Double Burden of Under- and Overnutrition and Nutrient Adequacy among Chinese Preschool and School-aged Children in 2009-2011.” European Journal of Clinical Nutrition, 69(12): 1323–1329.

Rangel, Marcos A. 2006. “Alimony Rights and Intrahousehold Allocation of Resources: Evidence from Brazil.” Economic Journal, 116(513): 627–658.

Silverstein, Merril, and Wencheng Zhang. 2020. “Grandparents’ Financial Contributions to Grandchildren in Rural China: The Role of Remittances, Household Structure, and Patrilineal Culture.” The Journals of Gerontology: Series B, 75(5): 1042–1052.

Song, Yang. 2014. “What Should Economists Know about the Current Chinese Hukou System?.” China Economic Review, 29 200–212.

Song, Yi, Jun Ma, Hai Jun Wang, Zhiqiang Wang, Peijin Hu, Bing Zhang, and Anette Agard. 2015. “Secular Trends of Obesity Prevalence in Chinese Children from 1985 to 2010: Urban-Rural Disparity.” Obesity, 23(2): 448–453.

Wells, Jonathan C, Ana Lydia Sawaya, Rasmus Wibaek, Martha Mwangome, Marios S Poullas, Chittaranjan S Yajnik, and Alessandro Demaio. 2020. “The double burden of malnutrition: aetiological pathways and consequences for health.” The Lancet, 395(10217): 75–88.

Yang, Jinyang, and Muchin I.A Bazan Ruiz. 2021. “Are pilot experiments random? Social connections and policy expansion in China.” The Journal of the Economics of Ageing, 18, p. 100305.

Zhang, Linxiu, Max Kleiman-Weiner, Renfu Luo, Yaojiang Shi, Reynaldo Martorell, Alexis Medina, and Scott Rozelle. 2013. “Multiple Micronutrient Supplementation Reduces Anemia and Anxiety in Rural China’s Elementary School Children.” The Journal of Nutrition, 143(5): 640–647.

